# Human Genetics of Face Recognition: Discovery of *MCTP2* Mutations in Humans with Face Blindness (Congenital Prosopagnosia)

**DOI:** 10.1101/2021.09.27.21263237

**Authors:** Yun Sun, Weiwei Men, Wan Fang, Enxing Zhou, Wei Yang, Zhiqiang Li, Hou-Feng Zheng, Yi Rao

## Abstract

Face recognition is important for both visual and social cognition. While congenital prosopagnosia (CP) or face blindness has been known for seven decades and electrophysiological studies have characterized face specific neurons for half a century, no molecular analyses have been undertaken. Here we report results of research combining classic genetics and modern genomics. From a large family with 18 CP members, we uncovered a fully co-segregating private mutation in the *MCTP2* gene which encodes a calcium binding transmembrane protein expressed in the central nervous system. More rare mutations in *MCTP2* were detected in CP families and were also associated with CPs in the cohort study. In another cohort of 1757, face recognition was different between 14 carriers with a frameshift mutation S80fs in *MCTP2* and 19 non-carrying volunteers. 6 families including one with 10 members showed the S80fs-CP correlation. Functional magnetic resonance imaging (fMRI) indicates that impaired recognition of individual faces by CPs with the *MCTP2* mutations is associated with inability to recognize the same faces in the right fusiform face area (rFFA). Our results have revealed the genetic predisposition of *MCTP2* mutations in CP, 74 years after the initial report of CP. This is the first time a gene required for a higher form of visual social cognition was found in humans.

## INTRDUCTION

Although human cognition is fascinating, molecular studies of human cognition are rare. Often, genes were first identified in animals before their studies in humans were undertaken. This approach limits the phenotype to those present in animals, often lower animals because it is difficult to uncover genes by function in non-human primates. Thus, it is difficult to study cognition existing only in humans or those not present in lower animals. However, genetic studies of human diseases have been highly successful ^1–3^. It is unnecessary to assume that human cognition is fundamentally different from diseases in amenibility to genetic studies. Genetics provides a non-invasive approach to study human cognition. Over the last decade, we have carried out several genome-wide association studies (GWAS) of human cognition from memory, social conformity to perceptual switching and top-down control in visual cognition ^4–8^. In spite of the findings of associated markers, we do not know whether the genes harboring or around the markers are causally linked. Genetically, linkage analyses in large families have succeeded in identifying human disease causing genes. We have undertaken this approach to investigate the molecular genetic basis of face recognition and results from 7004 subjects are reported in this paper.

Face recognition, one of the highest forms of visual cognition, is essential for social cognition ^9–12^. Neurons responding specifically to the face have been discovered in the interotemporal cortex (IT) ^13–22^ and the superior temporal sulcus (STS) ^13^ of monkeys for nearly half a century. Electric stimulation in monkeys has functionally implicated neurons in face recognition ^23; 24^. Face specific neurons have also been detected by event-related potentials ^25–27^ or direct electrophysiological recordings ^28^ in humans. Face responding brain areas have been detected in humans with positron emission tomography (PET) ^29; 30^ and functional magnetic resonance imaging (fMRI) ^31–41^. Results from trancranial magnetic stimulation are consistent with these regions being functionally important for face recognition ^42–44^.

Congenital prosopagnosia (CP), also known as developmental prosopagnosia, hereditary prosopagnosia (OMIM 610382), first reported in 1947 ^45^, is a selective impairment of visual learning and recognition of faces, in the absence of any detectable neurological injury ^46–53^. Both a questionnaire-based screening method ^54–56^ and behavioral tests ^57^ have estimated the prevalence of CP at 1.8 to 2.9% in the general population, providing a global estimate of millions of CP individuals (CPs). Familial ^46; 54; 55; 58–65^ and twin studies of CP ^42; 66–68^ suggest that CP and face recognition abilities are highly heritable. In pedigree analysis, a simple autosomal dominant mode of inheritance has often been observed ^54; 58; 60; 62; 64; 65^, indicating that mutations in a single gene can lead to difficulties in face recognition.

Beginning with a large pedigree including 18 members with obvious difficulties in face recognition in daily life, we have discovered a CP susceptibility gene, multiple C2 domains, transmembrane 2 (*MCTP2,* NM_018349). More rare mutations in *MCTP2* and correlations in families were detected in people at the lower end with 2 standard deviations (SDs) below the mean by an expanded screening from a cohort of 2904 individuals. In a second cohort of 1928, correlation of the rare alleles in *MCTP2* with the face recognition ability was detected in males by a gene based association analysis. Through a reverse-phenotyping approach, qualitative differences in face recognition were detected between 14 carriers with the same frameshift mutation S80fs in *MCTP2* and 19 non-carrying volunteers from a third cohort of 1757. From the 14 carriers, 6 families including one with 10 members also showed genotype-phenotype correlation. fMRI results indicate that impaired recognition of individual faces by CPs with the *MCTP2* mutations is associated with abnormal responses to repeated faces of the same identity in the right fusiform face area (rFFA). Our discovery may stimulate further research using genetics and genomics to study higher cognition in humans.

## RESULTS

### A Three-Generation Pedigree with CP

35 members from a three-generation pedigree (family A) (Table 1, Table S1, Figure 1a), were examined by an interview-based method developed previously ^51; 54; 60^ and used by many research groups ^69–79^. The mean age at interview was 42.5 ± 18.7 (SD) years (range of 15-78 years). 18 family members showed obvious difficulties in face recognition in daily life: 9 individuals (IV:2, IV:4, V:9, V:10, V:11, V15, VI:5, VI:6 and VI:9) had been aware of this deficit in their early life before we contacted them; 4 individuals (IV:6, V:4, V:6 and VI:8) had difficulties in recognizing people for a long time without knowing the reason; while the remaining 5 individuals (IV:10, V:1, V:13, V:19 and VI:7) who ignored this problem in their daily life, adopted strategies relying on cues other than the face (Table 1). There was significantly negative correlation between the spontaneous need of gaze contact and the experience of face recognition difficulty or the use of compensatory strategies (*p* < 0.001, r = - 0.658, n = 35). None of the persons showed signs of an autism spectrum disorder or neurodegenerative disorders.

**Figure 1:**
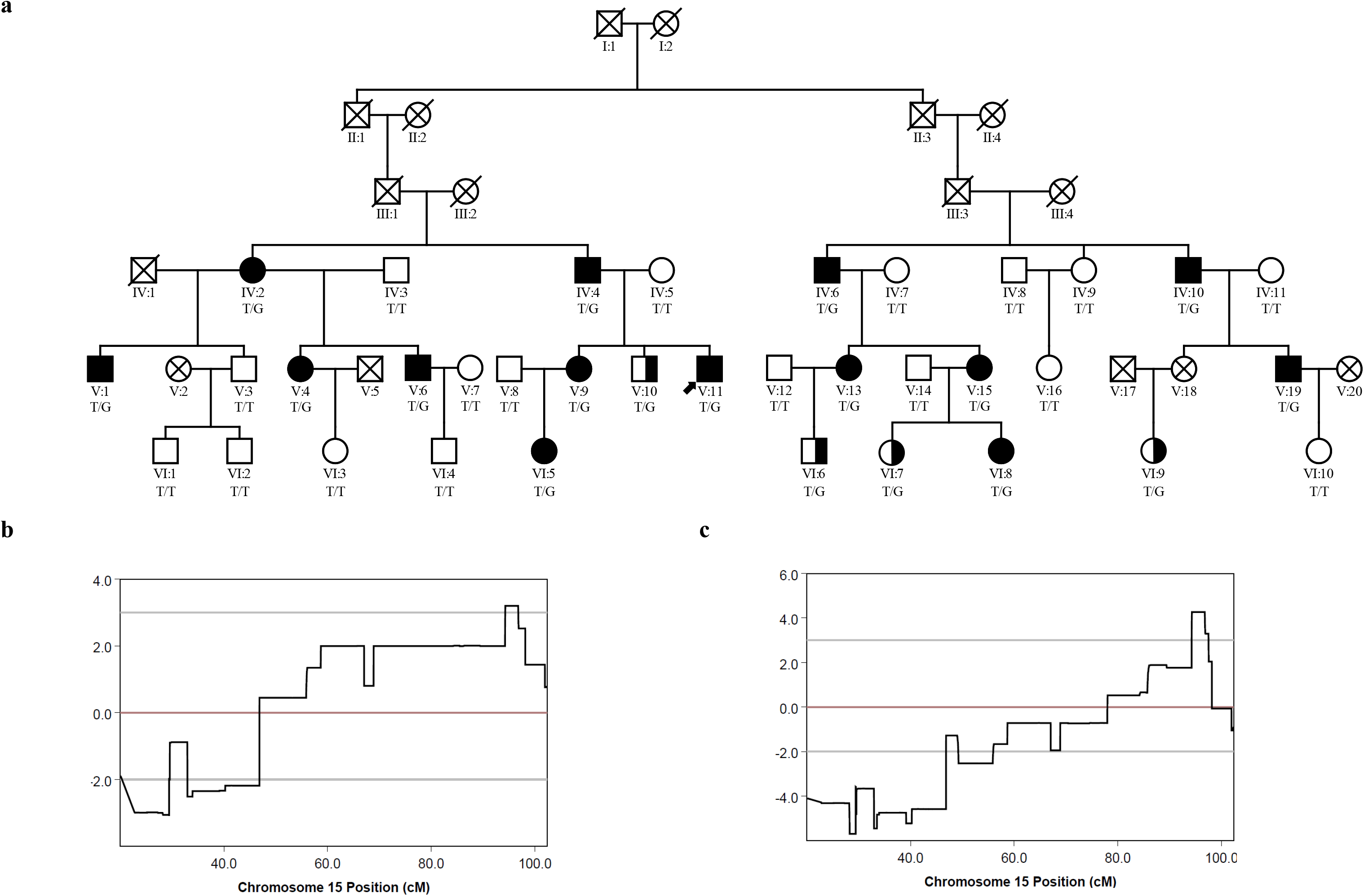
Mapping of a Novel Autosomal Dominant CP Locus to the Short Arm of Chromosome 15 (15q26.2). (a) The pedigree plot of family A with genotypes for c.T2147G, p.I716S, NM_018349 and phenotypes. Crosses, DNA samples not available; diagonal slashes, dead members; filled symbols, CP candidates in linkage 1; half-filled symbols, added as CP candidates in linkage 2; an arrow head, the index of family A. (b) Graphical representation of parametric linkage results of linkage 1 (LOD score = 3.20) in family A with diagnosis based only on interviews as well as behavioral tests, assuming a rare dominant model. (c) Graphical representation of parametric linkage results of linkage 2 (LOD score = 4.27) in family A with diagnosis based only on interviews but not behavioral tests, assuming a rare dominant model.

**Table 1.**
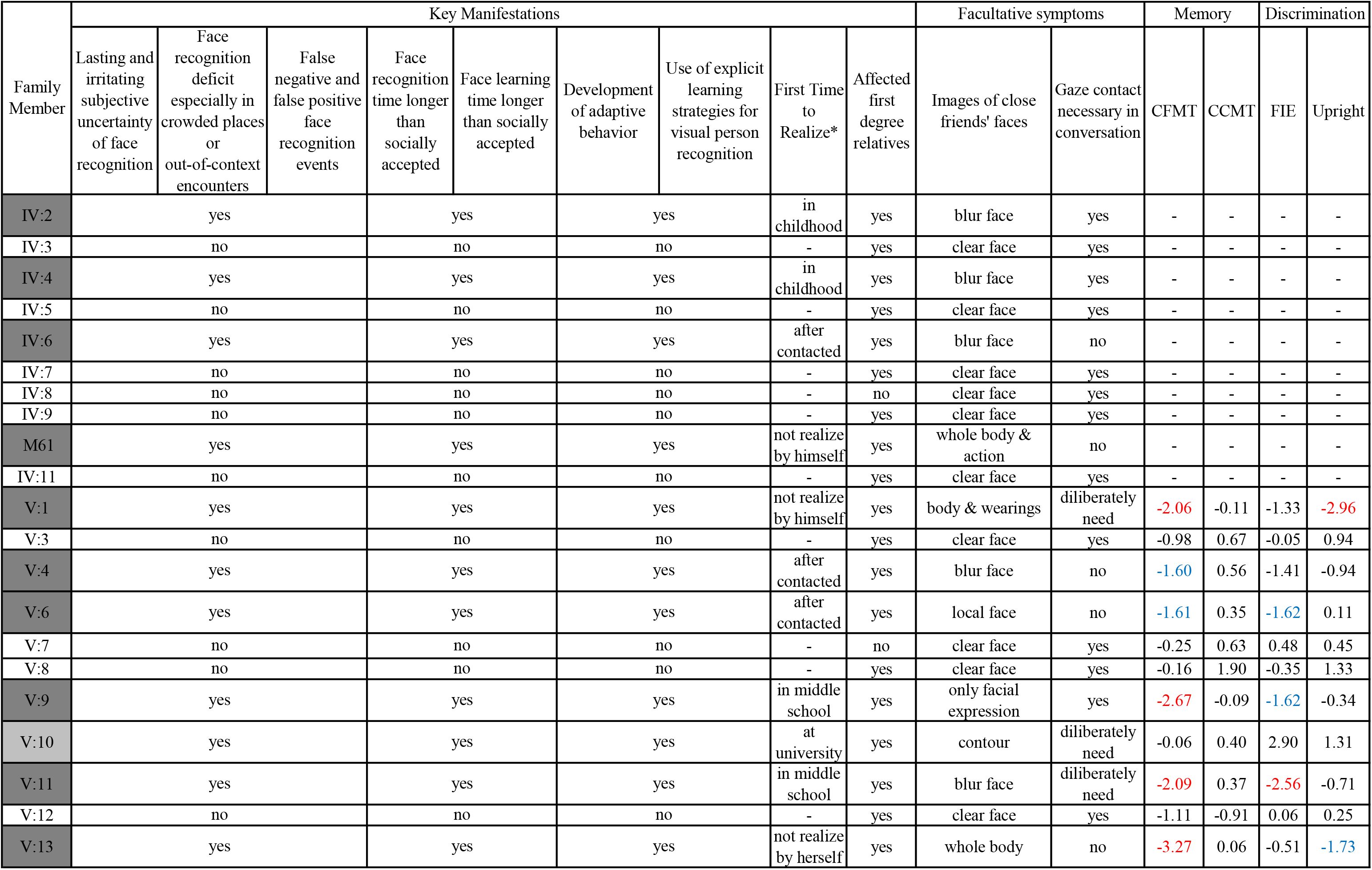
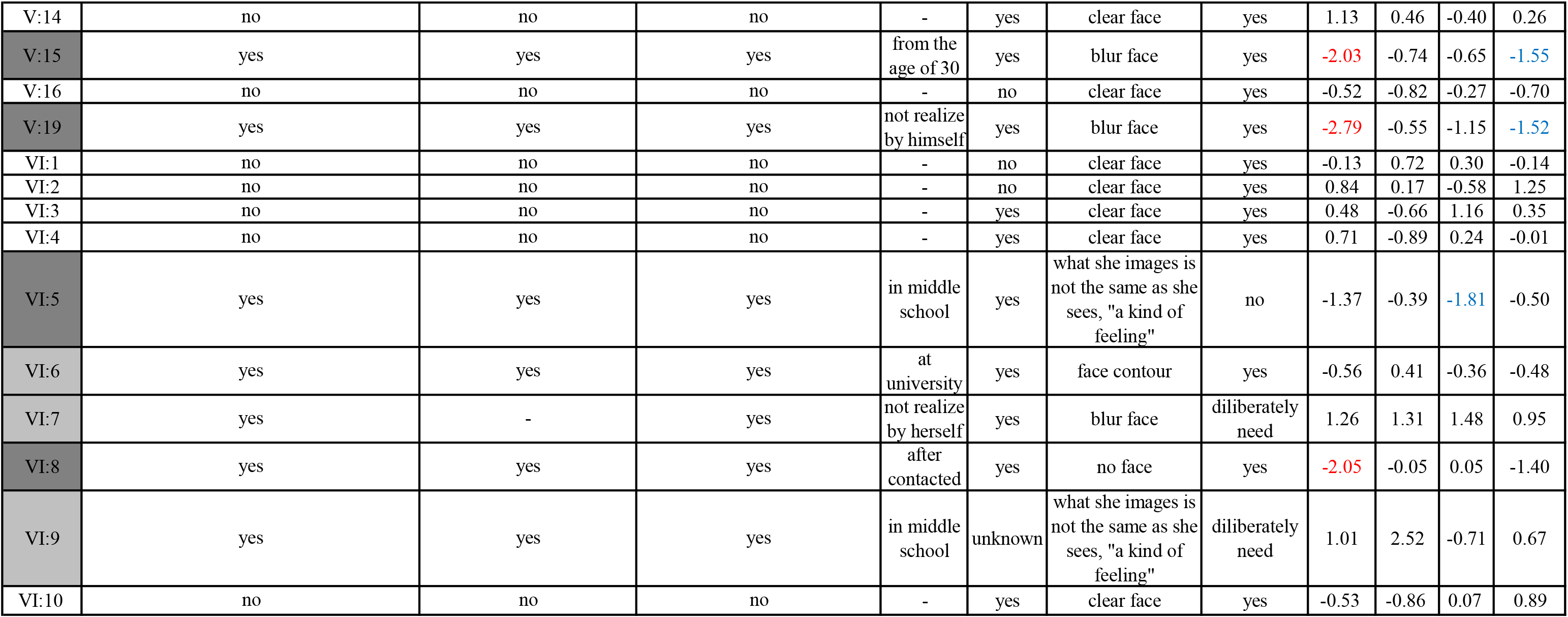
Diagnosis of CP in Family A. Members with dark grey background indicate CP candidates in linkage 1. Members with light grey background were added as CP candidates in linkage 2. Face processing z scores for members younger than 60 years of age in family A are coded as described in the method such that negative values indicate performances poorer than the mean. Red indicates below −2 SDs. Blue indicates below −1.5 SDs. Whether there is an obvious key manifestation for CP is expressed by yes or no. More detailed diagnostic information can be obtained by contacting the corresponding author.

| Family Member | Key Manifestations |  |  |  |  |  |  |  |  | Facultative symptoms |  | Memory |  | Discrimination |  |
| --- | --- | --- | --- | --- | --- | --- | --- | --- | --- | --- | --- | --- | --- | --- | --- |
|  | Lasting and irritating subjective uncertainty of face recognition | Face recognition deficit especially in crowded places or out-of-context encounters | False negative and false positive face recognition events | Face recognition time longer than socially accepted | Face learning time longer than socially accepted | Development of adaptive behavior | Use of explicit learning strategies for visual person recognition | First Time to Realize* | Affected first degree relatives | Images of close friends' faces | Gaze contact necessary in conversation | CFMT | CCMT | FIE | Upright |
| IV:2 | yes |  |  | yes |  | yes |  | in childhood | yes | blur face | yes | - | - | - | - |
| IV:3 | no |  |  | no |  | no |  | - | yes | clear face | yes | - | - | - | - |
| IV:4 | yes |  |  | yes |  | yes |  | in childhood | yes | blur face | yes | - | - | - | - |
| IV:5 | no |  |  | no |  | no |  | - | yes | clear face | yes | - | - | - | - |
| IV:6 | yes |  |  | yes |  | yes |  | after contacted | yes | blur face | no | - | - | - | - |
| IV:7 | no |  |  | no |  | no |  | - | yes | clear face | yes | - | - | - | - |
| IV:8 | no |  |  | no |  | no |  | - | no | clear face | yes | - | - | - | - |
| IV:9 | no |  |  | no |  | no |  | - | yes | clear face | yes | - | - | - | - |
| M61 | yes |  |  | yes |  | yes |  | not realize by himself | yes | whole body & action | no | - | - | - | - |
| IV:11 | no |  |  | no |  | no |  | - | yes | clear face | yes | - | - | - | - |
| V:1 | yes |  |  | yes |  | yes |  | not realize by himself | yes | body & wearings | diliberately need | -2.06 | -0.11 | -1.33 | -2.96 |
| V:3 | no |  |  | no |  | no |  | - | yes | clear face | yes | -0.98 | 0.67 | -0.05 | 0.94 |
| V:4 | yes |  |  | yes |  | yes |  | after contacted | yes | blur face | no | -1.60 | 0.56 | -1.41 | -0.94 |
| V:6 | yes |  |  | yes |  | yes |  | after contacted | yes | local face | no | -1.61 | 0.35 | -1.62 | 0.11 |
| V:7 | no |  |  | no |  | no |  | - | no | clear face | yes | -0.25 | 0.63 | 0.48 | 0.45 |
| V:8 | no |  |  | no |  | no |  | - | yes | clear face | yes | -0.16 | 1.90 | -0.35 | 1.33 |
| V:9 | yes |  |  | yes |  | yes |  | in middle school | yes | only facial expression | yes | -2.67 | -0.09 | -1.62 | -0.34 |
| V:10 | yes |  |  | yes |  | yes |  | at university | yes | contour | diliberately need | -0.06 | 0.40 | 2.90 | 1.31 |
| V:11 | yes |  |  | yes |  | yes |  | in middle school | yes | blur face | diliberately need | -2.09 | 0.37 | -2.56 | -0.71 |
| V:12 | no |  |  | no |  | no |  | - | yes | clear face | yes | -1.11 | -0.91 | 0.06 | 0.25 |
| V:13 | yes |  |  | yes |  | yes |  | not realize by herself | yes | whole body | no | -3.27 | 0.06 | -0.51 | -1.73 |

|  |  |  |  |  |  |  |  |  |  |  |  |
| --- | --- | --- | --- | --- | --- | --- | --- | --- | --- | --- | --- |
| V:14 | no | no | no | - | yes | clear face | yes | 1.13 | 0.46 | -0.40 | 0.26 |
| V:15 | yes | yes | yes | from the age of 30 | yes | blur face | yes | -2.03 | -0.74 | -0.65 | -1.55 |
| V:16 | no | no | no | - | no | clear face | yes | -0.52 | -0.82 | -0.27 | -0.70 |
| V:19 | yes | yes | yes | not realize by himself | yes | blur face | yes | -2.79 | -0.55 | -1.15 | -1.52 |
| VI:1 | no | no | no | - | no | clear face | yes | -0.13 | 0.72 | 0.30 | -0.14 |
| VI:2 | no | no | no | - | no | clear face | yes | 0.84 | 0.17 | -0.58 | 1.25 |
| VI:3 | no | no | no | - | yes | clear face | yes | 0.48 | -0.66 | 1.16 | 0.35 |
| VI:4 | no | no | no | - | yes | clear face | yes | 0.71 | -0.89 | 0.24 | -0.01 |
| VI:5 | yes | yes | yes | in middle school | yes | what she images is not the same as she sees, "a kind of feeling" | no | -1.37 | -0.39 | -1.81 | -0.50 |
| VI:6 | yes | yes | yes | at university | yes | face contour | yes | -0.56 | 0.41 | -0.36 | -0.48 |
| VI:7 | yes | - | yes | not realize by herself | yes | blur face | diliberately need | 1.26 | 1.31 | 1.48 | 0.95 |
| VI:8 | yes | yes | yes | after contacted | yes | no face | yes | -2.05 | -0.05 | 0.05 | -1.40 |
| VI:9 | yes | yes | yes | in middle school | unknown | what she images is not the same as she sees, "a kind of feeling" | diliberately need | 1.01 | 2.52 | -0.71 | 0.67 |
| VI:10 | no | no | no | - | yes | clear face | yes | -0.53 | -0.86 | 0.07 | 0.89 |

We further investigated face recognition among 25 family members younger than 60 by the Cambridge Face Memory Test-Chinese (CFMT-C) ^57; 80^, the matched Cambridge Car Memory Test (CCMT) ^81^ and the Face Inversion Effect (FIE) Discrimination test ^82^. Consistent with previous studies ^61; 62^, there was intra-familial heterogeneity in face recognition. 14 members reported experienced substantial real-world face recognition difficulties, with 10 individuals found to be impaired on at least one of the face behavioral tests with a Z value below 1.5 SDs. V:1, V:9, V:11, V:13, V:15, V:19 and VI:8 had a score more than two SDs below the normal level on CFMT-C, and V:4 (z = −1.60) and V:6 (z = −1.61), performed noticeably poorly (under 1.5 SDs). In the FIE discrimination test for holistic processing, the face-inversion effect of V:6 (z = −1.62), V:9 (z = −1.62), V:11 (z = −2.56) and VI:5 (z = −1.81) was impaired. V:1 (z = −2.96), V:13 (z = −1.73), V:15 (z = −1.55) and V:19 (z = −1.52) also performed poorly on the ability of discriminating upright faces. There were four individuals (V:10, VI:6, VI:7 and VI:9) who reported everyday difficulties but performed at normal levels on the tests of face recognition. All of them performed well in the CCMT test, indicating that their recognition of other physical stimuli was normal and independent of their abilities to recognize faces. Z scores for each member were listed in Table 1.

### A Specific Region on Chromosome 15q Containing a Candidate CP Susceptibility Gene

To position the CP susceptibility gene, we identified 10 members (V:1, V:4, V:6, V:9, V:11, V:13, V:15, V:19, VI:5 and VI:8) as CP cases. They were younger than 60, repeatedly had difficulties in recognizing faces in everyday life via interview and showed impairments in at least one of the face behavioral tests. 4 members (IV:2, IV:4, IV:6 and IV:10) were also CPs with lifelong difficulties in face recognition, though they were over 60 and not suitable for behavioral tests. To be cautious, those with normal performance on behavioral tests but obvious face recognition failure experience (V:10, VI:6 and VI:7) were classified as unclear. Remaining 17 family members (IV:3, IV:5, IV:7, IV:8, IV:9, IV:11, V:3, V:7, V:8, V:12, V:14, V:16, VI:1, VI:2, VI:3, VI:4 and VI:10) were classified as normal for the following genome-wide linkage analysis (named linkage 1) (Table S1).

Parametric linkage analyses with 177,126 SNPs (LD threshold r^2^ < 0.5) were performed, assuming an autosomal dominant inheritance with 80% penetrance, disease allele frequency at 0.001, and phenocopy rate at 0.05. Results of the genome-wide screen are shown in Figure S1a. The highest LOD (logarithm of odds) score 3.20, which suggested a possible CP-linked gene on chromosome 15, spanned 2.5 megabases (Mb) from rs6497118 (chr15:94266567, 15q26.1) to rs1437588 (chr15:96809782, 15q26.2) (Figure 1b). Keeping the pedigree structure, computer simulations led to an empirical *p* value of 0.017 suggesting that this result was significant at the genome-wide level (Table S2). Four other regions 2p25.1-24.3, 6q16.3, 12p12.1 and14q12 had LOD scores exceeding 1.0 (Figure S1a), but did not reach genome-wide significance by computer simulations (Table S2). Haplotype analysis was used for error elimination during the linkage scan. For the minimum candidate region (MCR), we constructed a genetic haplotype that was shared by all 14 affected individuals but not by the 17 unaffected individuals using markers with LOD scores over 3 (Figure S2a).

It is possible that some affected persons completed face recognition tasks with coping strategies for individual recognition without face recognition, leading to ambiguities in classifications ^60; 83; 84^. Thus, we did a second linkage analysis (linkage 2) after adding three individuals (V:10, VI:6 and VI:7), who were set as unclear in the linkage analysis analyzed above (linkage 1), also as CPs (Table S1). Linkage 2 extended results of linkage 1 in further supporting the region between rs6497114 (chr15:94245722) and rs12900387 (chr15:97520230) with LOD scores over 3 (maximum LOD score = 4.27), spanning 3.27 Mb on chromosome 15q26.1-26.2 (Figure 1c, Figure S1b). Two other regions 10q25.1-25.2 and 12p12.1 had LOD scores over 1.0. Keeping the pedigree structure, simulations were performed to assess empirical *p* values with 1,000 replicates to exclude false positive results due to random chances, and a significant *p* < 0.009 for the maximum LOD score on chr15 existed (Table S2). A genetic haplotype shared by all 17 affected but not by the 17 unaffected individuals using markers with LOD scores over 3, narrowed the MCR to a 1 Mb interval between rs1573086 and kgp9835458 (Figure S2b).

Candidate genes annotated by the newest NCBI database (https://www.ncbi.nlm.nih.gov/genome/gdv/) in the regions of both linkage analyses with LOD scores over 3 are listed in Table S3.

Copy number variations (CNVs) were identified for each member. No correlation was detected between the CNV genotypes and the phenotypes for each linkage analysis (data available upon request).

### A Mutation in *MCTP2* Revealed by Whole-Exome Sequencing

To find causative mutations on chromosome 15q26.1-26.2 linked to CP in family A, we employed the whole-exome sequencing (WES) approach with four affected individuals (V:6, V:9, V:11, and V:19, Table S1). After the alignment and variant calling, the mean depth of all samples was 83.55, and 94.6% of the targeted bases were covered at more than 10-fold on average (Table S4). In linkage regions with LOD scores over 3, 100% coding bases were covered at more than 20-fold in at least one of the four individuals. We excluded variants with minor allele frequency (MAF) > 0.05 in multiple databases, including the dbSNP (v150) ^85^, the 1000 Genomes Project (1000G) ^86–88^, the Exome Aggregation Consortium (ExAC) ^89^ and the Genome Aggregation (genomAD, http://gnomad.broadinstitute.org) databases. In the linkage region with LOD scores over 3, only one variant (c.T2147G; p.I716S, NM_018349) in *MCTP2* at chr15:94983466, was heterozygous in all the four CPs. This missense mutation (c.T2147G, p.I716S) was private to family A, not present in the dbSNP (v150), 1000G, ExAC and genomAD databases, albeit at this location another SNP rs200314451 (c.T2147C; p.I716T) had been reported with a MAF of 2.471e-05 (ExAC_ALL) in the ExAC database, with a MAF of 2e-04 in the 1000G, but not present in genomAD. In our expanded cohorts of 3,600 Chinese samples, we did not find the same mutation.

To answer whether this variant is a strong candidate of CP predisposition, we performed direct Sanger sequencing (primer sequences in Table S5) and co-segregation analysis in the pedigree. We found that the mutation (p.I716S) fully co-transmitted over multiple generations with all 18 family members who had difficulties in recognizing faces in daily life, including those with poor performance on the face behavioral tests and those with normal performance, but not in other family members (Figure 1a). We also performed the whole-genome sequencing (WGS) in two distantly related affected individuals V:19 and VI:5. We filtered our WGS datasets to exclude all variants that have a MAF over 0.05 in the dbSNP, 1000G, Exac and genomAD databases, and filtered for variants within the linkage regions with LOD scores over 3 by all affected individuals. Using this approach, 11 heterozygous variants detected in the linkage region were confirmed by Sanger sequencing (Tables S6 and S7). Notably, only one exonic variant (c.T2147G, p.I716S) occurred within Exon 17 (hg19, NM_018349) of *MCTP2*, as the result from WES. Furthermore, for the structure variations in the MCR detected from the 2 individuals by WGS, we did not find any co-segregating in the family members. Thus, the *MCTP2* mutation encoding p.I716S seems to be the only functional variant shared in the MCR by all patients in this large CP family, and is predicted to be disease causing by SIFT ^90^.

The full length of the *MCTP2* gene spans 180 kilobases (kb) of genomic DNA, with 22 coding exons, encoding a protein with 878 amino acid residues separated into three C2 domains and two transmembrane regions (TMRs) (Figure S3a). I716, located in the first TMR, is highly conserved with primate species (Figures S3a and S3b). The expression data of microarray and RNA-seq of two people (H0351.2001, H0351.2002) from the Allen Human Brain Atlas mutually support MCTP2 expression in the right fusiform gyrus which contains the rFFA, the core brain region most consistently and robustly activated by the face selectively.

### Additional *MCTP2* Mutations in CPs and CP Families Identified by Expanded Screening

On the basis of the theoretical and experimental considerations, it has been suggested that rare functional alleles are important contributors to the genetics of phenotypes ^91–94^. The results from the study of a typical CP family A indicate that rare and even private, functional mutations in *MCTP2* could be related to a CP phenotype.

To examine whether the rare functional mutations in *MCTP2* were present in more CPs, we analyzed 2904 individuals by a questionnaire adapted from the 20-item self-report measure for quantifying prosopagnosia ^95^, similar to the interview we used for diagnosing family A. From these we identified 75 individuals who scored below 2 SDs of the mean among the 2904 participants and sequenced their *MCTP2* exons. Five rare heterozygous functional variants including one frameshift and four missense mutations were found in seven individuals (Figures 2a and 3a, Table S8).

**Figure 2:**
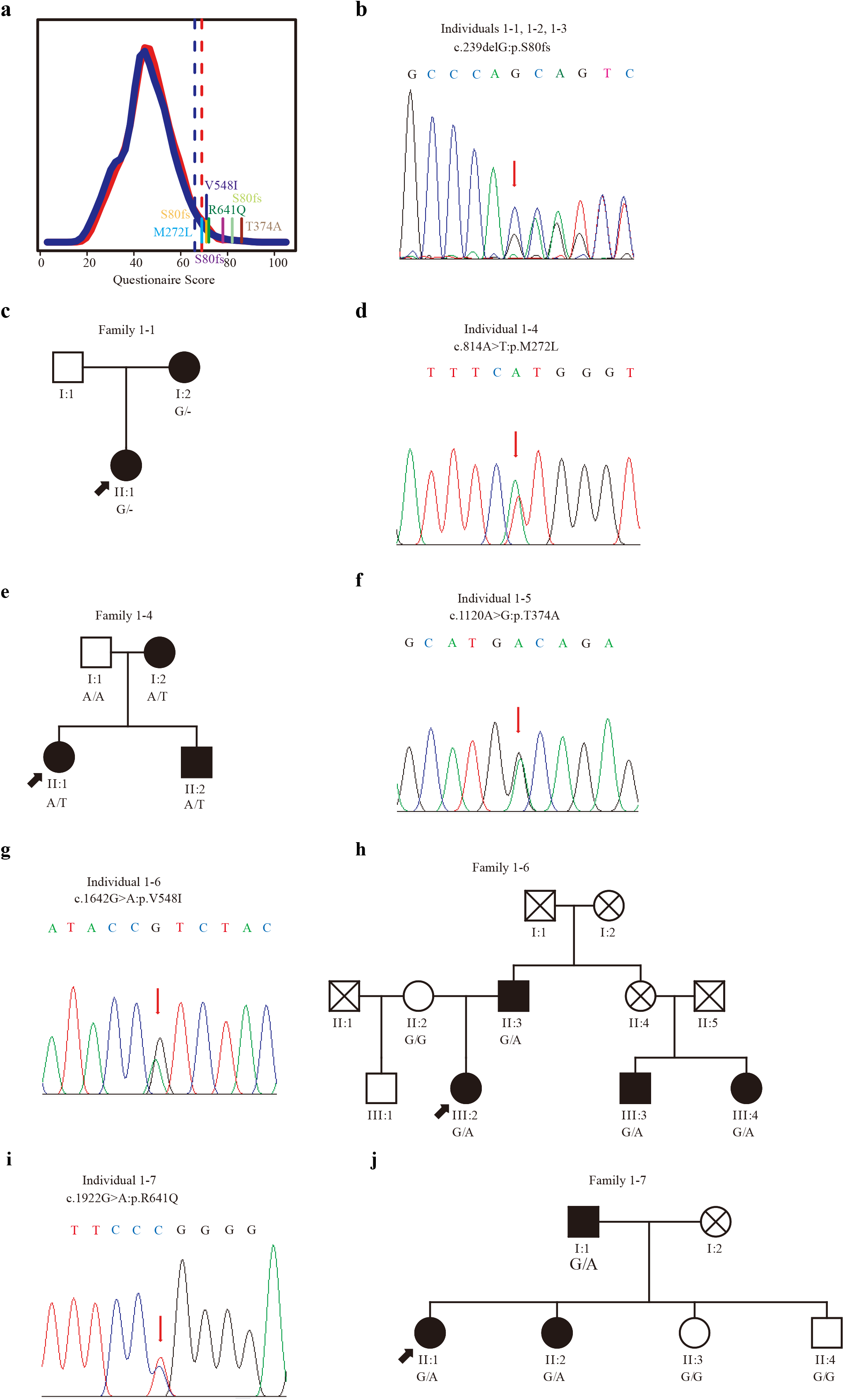
Mutations Detection in MCTP2 and Co-segregation in CP. (a) The distribution of the 20-item questionnaire scores in 743 males (the blue approximate normal curve) and 2161 females (the red approximate normal curve). Dashed lines indicate the cut-off of 2 SDs poorer than the mean. Sticks with different colors indicate different individuals with different mutations. (b) A mutation c.239delG, leading to p.S80fs in individuals 1-1, 1-2, 1-3 with CP. All three of them were aware of difficulties with face recognition early in their life (Table S9). (c) Correlation of the (c.239delG, p.S80fs) genotype with the CP phenotype in the family of 1-1, with 3 members tested. (d) A mutation c. 814A>T, leading to p. M272L in individual 1-4. (e) Correlation of the genotype (c.814A>T, p.M272L) with the CP phenotype in the family of 1-4. (f) A mutation c.1120A>G, leading to p.T374A in individual 1-5. (g) A mutation c.1642G>A, leading to p.V548I in individual 1-6. (h) Correlation of the (c.1642G>A, p.V548I) genotype with the CP phenotype in the family of 1-6, with 6 members tested (Table S9). (i) A mutation c.1922G>A, leading to p.R641Q in individual 1-7. (j) Correlation of the (c.1922G>A, p.R641Q) genotype with the CP phenotype in the family of 1-7, with 5 members tested (Table S9).

**Figure 3:**
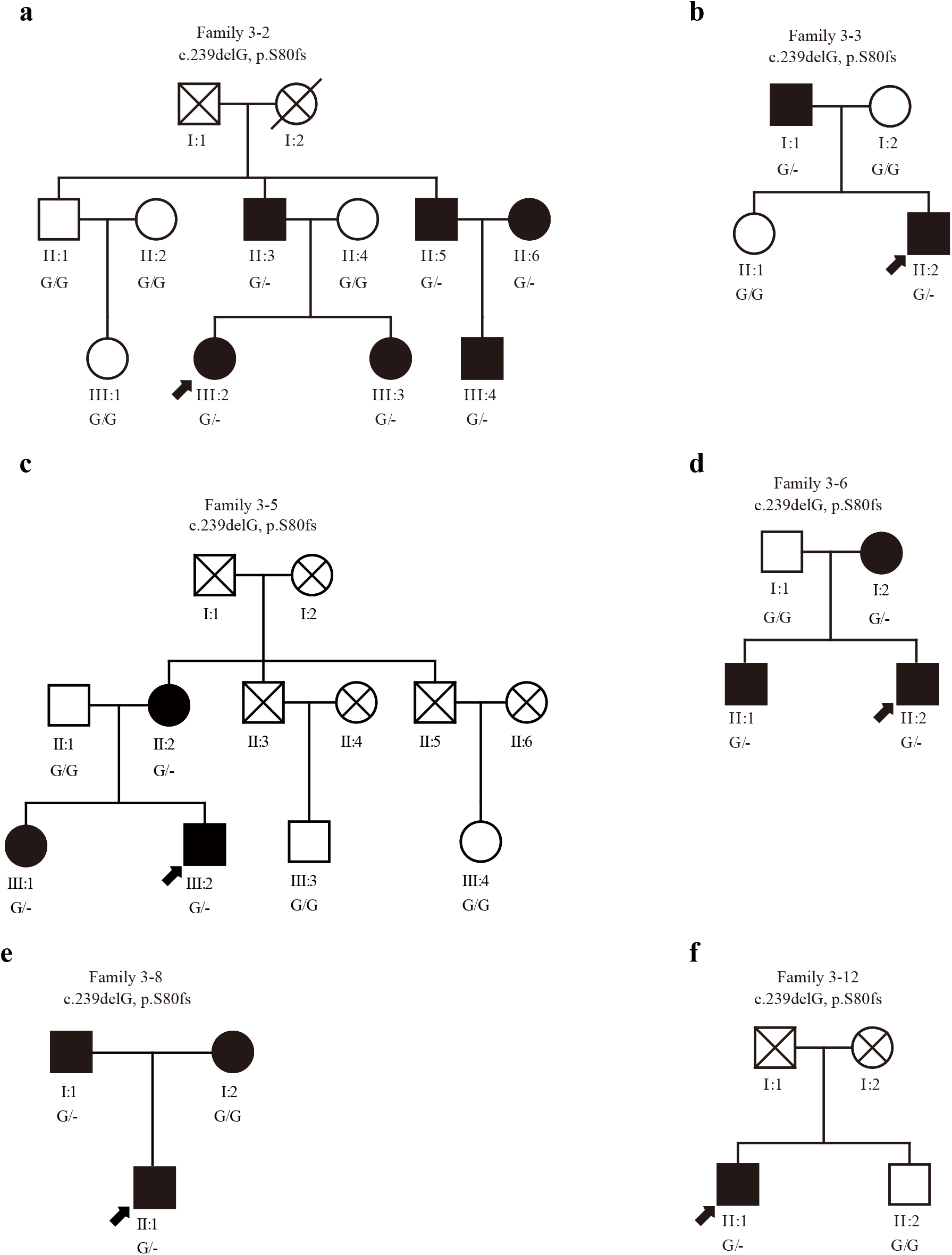
Co-segregation of c.239delG, p.S80fs in MCTP2 in CP. 16 frameshift deletion mutation (c.239delG, p.S80fs) were detected in a cohort of 1757 students. 14 of them were available for further analysis. More members from 6 families were available (Table S11). (a) Correlation of the (c.239delG, p.S80fs) genotype with the CP phenotype in the family of 3-2, with 10 family members tested. (b) Correlation of the (c.239delG, p.S80fs) genotype with the CP phenotype in the family of 3-3, with 4 family members tested. (c) Correlation of the (c.239delG, p.S80fs) genotype with the CP phenotype in the family of 3-5 with 6 family members tested. (d) Correlation of the (c.239delG, p.S80fs) genotype with the CP phenotype in the family of 3-6 with 4 family members tested. (e) Correlation of the (c.239delG, p.S80fs) genotype with the CP phenotype in the family of 3-8, with 3 members tested. (f) The pedigree plot of family 3-12 with genotype and phenotype.

The frameshift mutation (c.239delG, p.S80fs) in exon 1 of *MCTP2* was found in 3 of those 75 who reported poor performance in face recognition (individuals 1-1, 1-2 and 1-3) (Figure 2b). Further interviews revealed that all three individuals carrying p.S80fs reported lasting and irritating subjective uncertainties of face recognition, and realized this condition in their early life (Table S9). One relative of individual 1-1 carries p.S80fs and had adopted a strategy to observe other features, which took her more time to recognize a person (Figure 2c, Table S9).

In the family of individual 1-4, those who carry the p.M272L mutation in exon 5, reported their own strategies to recognize others, but still coped poorly with their difficulties (Figures 2d and 2e, Table S9). The p.T374A mutation in exon 8 was detected in individual 1-5 who was aware of having a severe deficit in face recognition (Figure 2f, Table S10). In the family of individual 1-6, the mutation p.V548I in exon 12 was correlated well with the phenotypes (Figures 2g and 2h, Table S9). In the family of individual 1-6, the mutation p.R641Q was found to segregate in all affected children of the family. These mutations are also evolutionarily conserved in animals (Figure S3b).

### A Link between Rare Alleles in *MCTP2* and Face Recognition Revealed by a Gene Based Association Analysis

In the next step, we further aimed to evaluate the association of the alleles in *MCTP2* that harbor rare coding variants of moderate or large effects on protein coding with face recognition abilities. We had a second cohort of 1928 individuals who took the same 20-item self-report measure as the first cohort of 2904 mentioned above. We resequenced all 22 exons, 5’UTR, 3’UTR and the exon-intron boundaries of *MCTP2* and did a single gene-based burden test for overall, female and male cohorts using the unified optimal sequence kernel association test (SKAT-O) ^96^.

After alignment and variant calling of all samples, 95% of targeted bases were covered at ≥ 10-fold per individual (Table S4). A total of 53 rare coding and four splice variants with MAF ≤ 0.005 were identified and verified by Sanger sequencing (Table S10). Five algorithms (Sift, Polyphen2_HDIV, Polyphen2_HVAR, LRT and Mutation Taster) integrated by ANNOVAR ^97^ were used to predict the functional severity of variants. Primary analyses based on ^98^ tested 1) nonsynonymous (NS) strict variants: disruptive variants (nonsense, essential splice site and frameshifts) plus missense variants predicted to be damaging by all five algorithms (22 individuals); 2) NSbroad variants: disruptive plus missense variants predicted to be damaging by at least one algorithm (71 individuals) and 3) NSall variants: disruptive plus all missense variants (124 individuals).

Significant association was detected with the disruptive variants plus all missense variants (NSall, *p* = 0.0266) in the overall cohort based on burden testing (rho = 1 as optimal). In the male cohort, there was a significant enrichment of rare variants associated with face recognition ability in all three categories by both burden testing and optimal testing: NSstrict (*p*_burden_ = 0.0113, *p*_optical_ = 0.0212), NSbroad (*p*_burden_ = 0.0029, *p*_optical_ = 0.0061) and NSall (*p*_burden_ = 0.0011, *p*_optical_ = 0.0024). No association was observed in the male cohort based on SKAT testing (rho = 0 as optimal). When considering the female cohort, no significant aggregation of rare variants was observed for each analysis. Details of the variants contributing to these significant test results are shown in Table 2. Our results suggest a high proportion of causal variants in *MCTP2* exert effects in the same direction and indicate the association between variant burden and face recognition in the male cohort, at least in part, by the effects of *MCTP2*.

**Table 2.** Gene Based Association Analysis for Rare Variants in MCTP2.

|  |  | All | Male | Female |
| --- | --- | --- | --- | --- |
| Nsstrict | Burden | 0.1286 | **0.0113** | 0.8746 |
|  | SKAT | 0.9560 | 0.7612 | 0.9009 |
|  | SKAT-O | 0.2249 | **0.0212** | 1.0000 |
| Nsbroad | Burden | 0.0603 | **0.0029** | 0.9518 |
|  | SKAT | 0.9914 | 0.7611 | 0.9276 |
|  | SKAT-O | 0.1141 | **0.0061** | 1.0000 |
| Nsall | Burden | **0.0266** | **0.0011** | 0.9229 |
|  | SKAT | 0.9368 | 0.7997 | 0.9744 |
|  | SKAT-O | 0.0499 | **0.0024** | 1.0000 |

### A Frameshift Mutation in CP Individuals

Results of pedigree and population studies support correlation between rare alleles in *MCTP2* changing protein coding with the ability of face recognition, implicating *MCTP2* in face recognition. As mentioned above, the frameshift deletion c.239delG (p.S80fs) in the first exon of *MCTP2* which would eliminate most of the MCTP2 protein was present in 3 of 75 individuals reporting poor face recognition from the first cohort of 2904 individuals. In our second cohort, the ExAC and the gnomAD databases in East Asian population, the MAF is 0.0026, 0.0031 and 0.0045 respectively. This frequency lies within the reported range of CP prevalence and is high enough to allow identification of additional carriers consenting to analysis of face recognition through a reverse-phenotyping approach.

We further screened in a third independent cohort of 1757 individuals for this frameshift mutation. We detected 16 individuals carrying this mutation in a heterozygous condition with a MAF of 0.0046. Among them, 14 individuals agreed to be further examined (Table S11). Our interviews documented qualitative differences in the behavior of daily face recognition between the 14 carriers and 19 non-carrying volunteers from the same cohort (Table 3).

**Table 3.** Differences in Daily Face Recognition between Target Group and Controls Regarding the p.S80fs Mutation.

| Manifestations of poor face recognition | Target group N=14<br>with the p.S80fs<br>mutation<br>(6 males/8 females) | Controls N=19<br>without the p.S80fs<br>mutation<br>(5 males/14 females) | Fisher's exact<br>test, p value, two-<br>tailed |
| --- | --- | --- | --- |
| Lasting and irritating subjective uncertainty<br>of face recognition | 9/14 | 1/19 | <0.001 |
| Face recognition deficit especially in<br>crowded places or<br>out-of-context encounters | 12/14 | 4/19 | <0.001 |
| False negative and false positive face<br>recognition events | 10/14 | 4/19 | 0.006 |
| Face recognition time longer than socially<br>accepted | 11/14 | 3/19 | <0.001 |
| Face learning time longer than socially<br>accepted | 11/14 | 3/19 | <0.001 |
| Development of adaptive behavior | 12/14 | 5/19 | 0.01 |
| Use of explicit learning strategies for visual<br>person recognition | 13/14 | 4/19 | <0.001 |
| Self-evaluation of impaired visual<br>recognition of faces | 5/14 | 1/19 | 0.062 |

Four individuals 3-1, 3-2, 3-3 and 3-4 had clearly realized their difficulties in face recognition before we contacted them. Examination of all available family members of 3-2 and 3-3 showed that this mutation segregated with face recognition deficits in both families (Figure 3a, Table S11).

Individual 3-5 did not report difficulties in recognizing people/faces, but he did feel different from others in the way of recognizing people/faces (Table S11). In the family of 3-5 (Figure 3c, Table S11), the mutation was also identified in two relatives (II:2 and III:1), who reported daily failures in face recognition.

For the remaining nine individuals during the interview, they thought they were the same as others or even better than others in face recognition (Table S11). However, eight individuals (from 3-6 to 3-13) developed adaptive behaviors and depended on explicit learning strategies for recognition, which made it not hard to recognize acquaintances (the whole person, not just the face) in their daily life. When they encountered strangers with few features, or actors especially actresses on the screen, or familiar people out-of-context, the strategies would not always work properly, but they could make adjustments and update the information quickly. The relatives of individual 3-6 who carry this mutation complained difficulties in recognition (Figure 3d, Table S11). For individual 3-8, the strategy to recognize people should be inherited from both parents (Figure 3e, Table S11). The relative of individual 3-12 did not show prosopagnosia and do not carry the mutation (Figure 3f, Table S11). One individual, 3-14 did not show obvious abnormal face recognition ability during the whole interview.

The inferred haplotypes around the c.239delG (p.S80fs) mutation were estimated by comparison to microsatellites data collected on mother-father-offspring trios (Figure S4) and excluded the possibility of the same founder origin.

### Neuroimaging Studies of Family CPs with the *MCTP2* Mutation

With the family A, we carried out fMRI analysis and compared results of three family members with CP and three without CP with normal controls. We found that impaired face recognition in CPs with the *MCTP2* mutation is associated with abnormal responses to individual faces in the fusiform face area (FFA).

The right FFA (rFFA) is the core brain region most consistently and robustly activated by the face selectively, and is involved in detecting the presence of a face and in discriminating individual faces ^33; 34; 99–104^.

We functionally localized the rFFA as the region of interest (ROI) in family members and a group of normal controls by contrasting the blood oxygenation level-dependent (BOLD) signal when participants viewed faces versus non-face stimuli. As in normal participants (Figure S5a), there were significant activities in the rFFA of family members with CP (A-VI:5, Figure S5b, A-VI:7, Figure S5c and A-VI:9, Figure S5d) and family members without CP (A-VI:1, Figure S5e, A-VI:2, Figure S5f and AVI:10, Figure S5g). These suggest that the rFFA of the CPs in family A could respond specifically to faces.

Another marker for the special face processing is the Face-Inversion Effect (FIE), to which the rFFA is also sensitive. Assessment of the FIE with the protocol embedded in the adaptation paradigm (Figures 4a, S6a) showed a significantly higher response to upright than to inverted faces in the rFFA in normal subjects (paired t-test *p* < 0.00001, Figure S6b). In addition, in every single control subject there was a higher activation level in response to the upright than the inverted faces: the contrast ‘upright face > inverted face’ was significant at *p* < 0.05 in 18/20 normal subjects and showed a non-significant trend in the predicted direction in the remaining subjects (*p* < 0.09, *p* < 0.14). Similar to normal controls, the non-CP members of A-VI:1 (Figure S6c), A-VI:2 (Figure S6d), A-VI:10 (Figure S6e) and CP members A-VI:5 (Figure S6f) and A-VI:9 (Figure S6h) showed a higher response to the upright than the inverted faces, except A-VI:7 who showed a lower response to the upright than the inverted faces (Figure S6g). The fMRI-FIE score (the percent signal change under the upright condition versus the inverted condition within the ROI) for each family member was within the range of the normal controls (Figure 5a), except the CP member A-VI:7 who showed a significantly different activation pattern compared with the normal controls (*p* = 0.0075, Figure 5a) using the Crawford and Howell’s modified t test ^105; 106^.

**Figure 4:**
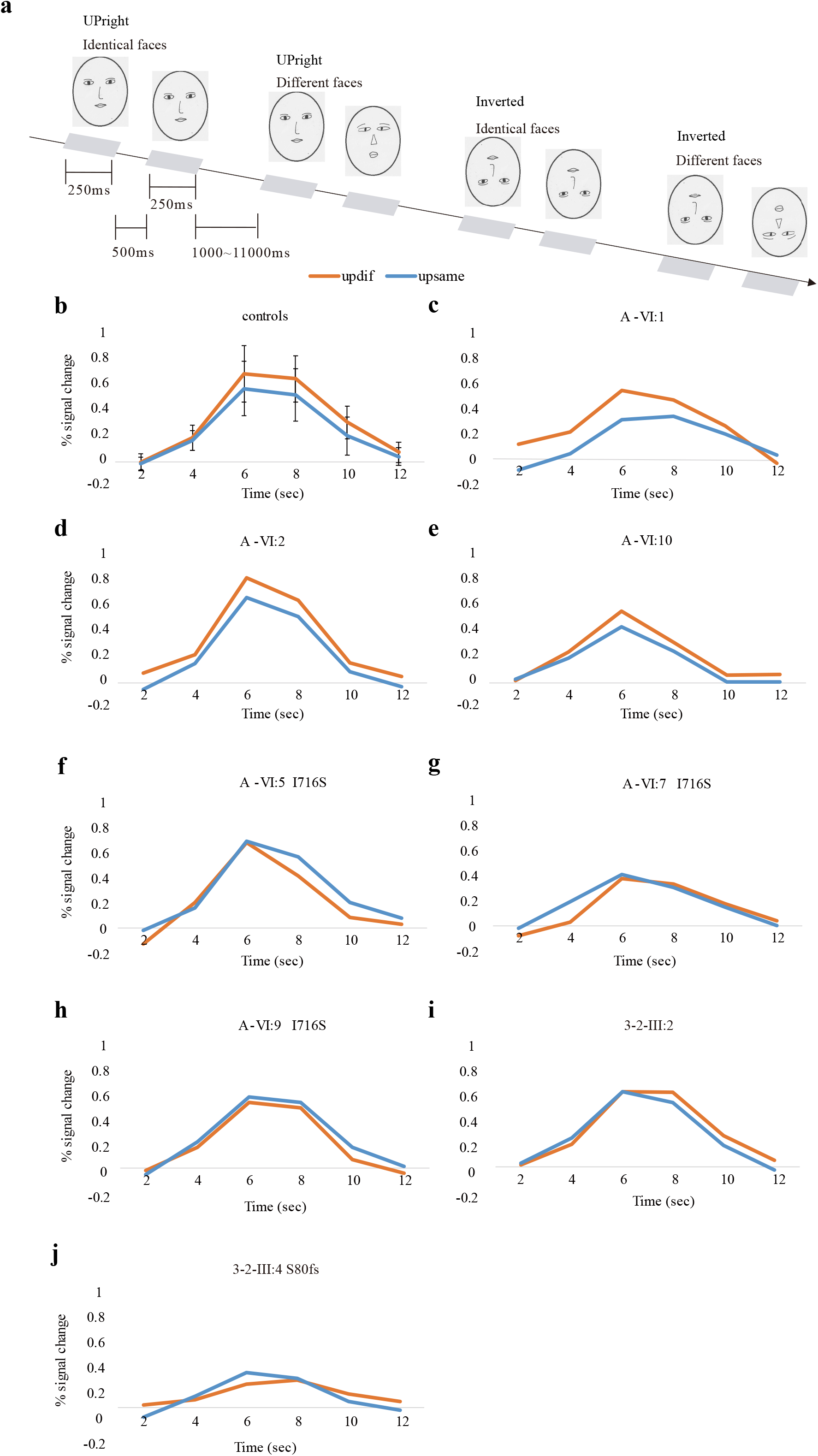
rFFA Activation in Face Discrimination. Experimental paradigm as Figure S6a. The average percent signal change (±SD for controls) from baseline fixation is plotted for the different and identical upright face conditions. Bold response to pairs of upright different faces or identical faces in the rFFA of controls (n=20) (b); non-CP members A-VI:1 (c), A-VI:2 (d), A-VI:10 (e) and CP members A-VI:5 (f), A-VI:7 (g), and A-VI:9 (h) from family A with the I716S mutation in MCTP2; non-CP 3-2-III:2 (i) and CP3-2-III:4 (j) from family 3-2 with the S80fs mutation in MCTP2.

**Figure 5:**
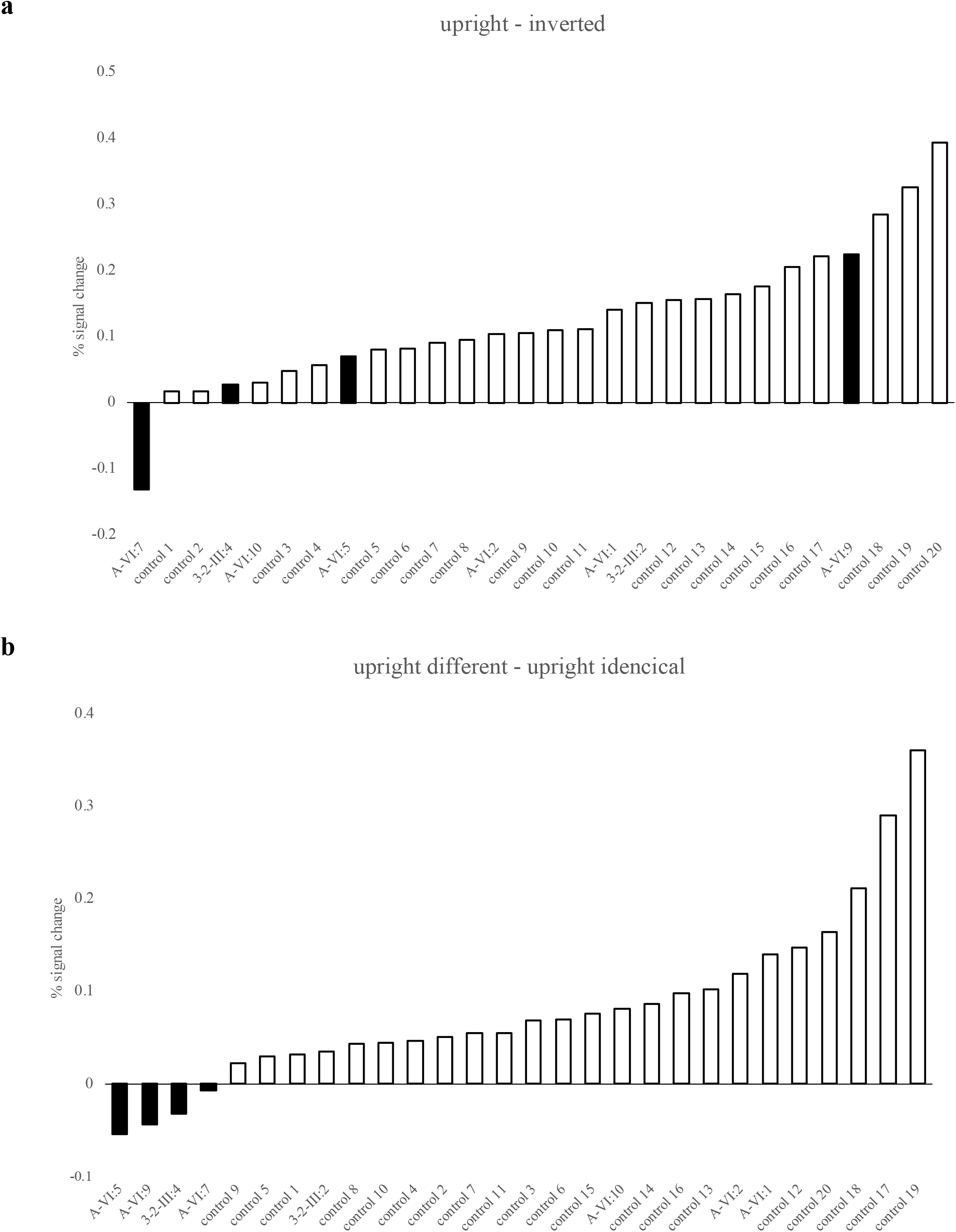
Comparison between CP and Control Individuals in the FIE and Adaptation Experiments. (a) The FIE score (difference in percent signal change between responses to upright different faces and inverted different faces) is plotted for CP individuals (black bars) and each individual control subject (white bars) in an increasing order. (b) The adaptation score (difference in percent signal change between responses to upright different faces and upright identical faces) is plotted for CP individuals (black bars) and each individual control subject (white bars) in an increasing order.

To test whether the rFFA of family CPs could process facial identities, we conducted experiments using an fMRI adaptation paradigm in an event-related design ^82^ with the above-defined ROI (Figure 4a). In group analysis of normal subjects, the expected repetition suppression from fMRI adaptation to facial identity was highly significant in the rFFA (paired t-test *p* < 0.0001, Figure 4b). In addition, in every single control subject there was a higher activation level in response to pairs of up different faces (UDF) than to pairs of up identical faces (UIF): the contrast ‘UDF>UIF’ was significant at *p* < 0.05 in 18/20 normal subjects and showed a non-significant trend in the predicted direction in the remaining subjects (*p* < 0.057, *p* < 0.25). In each family member without CP (A-VI:1, Figure 4c, A-VI:2, Figure 4d and A-VI:10, Figure 4e), there was a higher activation level in response to pairs of UDF than to pairs of UIF. The contrast ‘UDF>UIF’ was significant at *p* < 0.05 in each non-CP family member without the *MCTP2* mutation. In contrast, the fMRI signals in the rFFA of the CPs did not show larger response to different faces than identical faces (A-VI:5, Figure 4f, A-VI:7, Figure 4g and A-VI:9, Figure 4h). The region-specific attenuation of neuronal activity due to repetition of identical faces appeared to be reduced in CPs, while different faces yielded similar response in the rFFA in CPs, non-CPs and normal controls. The adaptation score (percent signal change under the upright different condition versus the upright identical condition within the ROI) was lower for CPs than for every individual control subject (Figure 5b) and there was a non-significant trend in the same direction for CPs compared to normal controls (A-VI:4, *p* = 0.05407, A-VI:6, *p* = 0.12832 and A-VI:8, *p* = 0.06548) using the Crawford and Howell’s modified t test ^105; 106^. These results show that rFFA responses to identical faces are impaired in CPs with the *MCTP2* mutation, implying a mechanism underlying the difficulty of famliy A CPs in identifying seen-before faces.

In the 3-2 family with the frameshift mutation in the 80^th^ amino acid in *MCTP2*, we observed a normal response to faces in rFFA at the categorical level both in CP III:4 (Figure S5h) and non-CP III:2 (Figure S5i), but a lack of FIE (Figure S6j) and a failure of repetition from fMRI adaptation to facial identity (Figure 4j) also occurred in the rFFA of CP III:4.

## DISCUSSION

Our findings indicate that mutations in the *MCTP2* gene contribute to CP in humans. This conclusion is supported by the following: 1) a CP locus at 15q26.1-q26.2 containing *MCTP2* was identified by linkage analysis in family A with autosomal dominant CP (Figure 1, Figure S1); 2) a private mutation c.T2147G (encoding p.I716S) in *MCTP2* was the only functional mutation identified by WES and WGS in the MCR that fully co-segregated with CP in the entire family A (Figure 1); 3) 5 rare functional mutations in *MCTP2* were found in 7 CP individuals from a group of 75 with poor face recognition out of a cohort of 2904 individuals (Figure 2); 4) of those 7 CPs, family members from 4 CPs were available for analysis and showed phenotype-genotype correlations (Figs.2c, 2e, 2h and 2j); 5) 3 out of 7 CP individuals from the cohort with 2904 subjects contained the frameshift deletion c.239delG, p.S80fs in the first exon of *MCTP2* (Figure 2b), a trio of one of these 3 CPs available for testing further supported the correlation between c.239delG, p.S80fs and CP (Figure 2c); 6) 16 individuals from another cohort of 1757 subjects contained the same frameshift deletion mutation c.239delG, p.S80fs, with 14 available for further analysis. Differences in the behavior of daily face recognition were detected between the 14 carriers and 19 non-carrying volunteers from the same cohort (Table 3); 7) four of 14 had unambiguous CP with two families available for analysis and all supported the correlation between c.239delG, p.S80fs and CP (Figures 3a and 3b), with further support from additional families (Figures 3c, 3d, 3e and 3f) who have developed explicit strategies to recognize people with non-facial clues to overcome their difficulties in face recognition; 8) correlation between the rare alleles in *MCTP2* and face recognition ability was detected in males by a gene based association analysis in a cohort of 1928 individuals. 9) impaired face recognition in CPs with the *MCTP2* mutations was associated with abnormal response to individual faces in the rFFA by neuroimaging studies.

*MCTP2* is not the only gene involved in CP. Only 7 of the 75 CPs from the 2904 cohort carried *MCTP2* mutations. Rare alleles in *MCTP2* were not correlated with face recognition ability in females from the cohort of 1928. More linkage studies would be helpful. Higher cognition involves many cells and molecules. Hypothesis-free genomic analysis should be reconsidered with the next generation sequencing method covering both common and rare genetic variants through larger sample sizes, exceeding a million human participants, to find more genetic clues and replicate our *MCTP2* results in human face recognition in the future.

### Molecular Studies of Face Recognition

Previous studies have been taken with behavioral, electrophysiological and genetic approaches, but no molecular studies have been undertaken.

In addition to primates ^9–11; 13; 20; 100^, face specific neurons have also been detected in sheep, dogs, chickens, pigeons ^107–111^.

In humans, differential responses to faces occur early in infancy ^112^ and responses to mother’s face occur within 4 to 35 days after birth ^113–121^.

There are humans defective in recognizing other objects but no face ^122–125^, and there are humans who have prosopagnosia, or face blindness, defective in recognizing faces but not other objects ^47; 126–132^. Acquired prosopagnosia includes damages by diseases or injuries ^132–139^. Several extended CP pedigrees have been reported ^61; 62; 65^, but the genes underlying their phenotype remain unclear.

We have performed GWAS of memory, social conformity, visual perceptual switching and visual top-down control in humans ^4–8^, and understand the limit of GWAS.

This paper represents the first molecular genetic study of face recognition. Our experience with GWAS of human cognition made us determined to use classic linkage analysis, rather than GWAS, to study genes involved in face recognition. An important lesson from our GWAS is that it is very difficult to study causal relationship from GWAS. We therefore persevered in using classic linkage analysis with genomic markers. The availability of family A is particularly helpful in allowing us to find an MCR with LOD score of 4.27 (Figure 1c). The discovery of *MCTP2* was confirmed in other CP individuals and trios or small families in China (Figures 2 and 3). We have found more families with other unknown genetic reasons and will further analyze them.

The expression data of microarray and RNA-seq of two people (H0351.2001, H0351.2002) from the Allen Human Brain Atlas support that *MCTP2* is expressed in the right fusiform gyrus, covering the face selective region. Although the expression level of *MCTP2* is relatively low, this does not prevent *MCTP2* from playing an important role.

The *MCTP2* gene encodes a protein with three C2 domains and two transmembrane regions (TMRs) with resemblance to proteins involved in synaptic transmission^140^. Its C2 domains can bind Ca^2+^^140^. Many proteins that bear the Ca^2+^-binding C2 domain are involved in membrane and vesicle trafficking, playing a central role in neuronal transmission^141; 142^. It has also been implicated in lipid droplets formation^143^.

The C2 domains and TMRs of MCTPs are evolutionarily conserved from invertebrates such as Caenorhabditis elegans and Drosophila melanogaster, that only have one *MCTP* gene, to mammals that have two genes (*MCTP1* and *MCTP2*). MCTP1 is expressed in the central nervous system and has been implicated in regulating endocytic recycling of specific CNS neurons and synapses^144^. In Drosophila, MCTP is involved in stabilizing synaptic transmission and homeostatic plasticity^145^. The four zebrafish *MCTP* genes are expressed mainly in the nervous and muscular systems. Knocking down *MCTP2b* impaired embryonic development^146^.

Both *MCTP2* and *MCTP1* had been associated with attention-deficit/hyperactivity disorder (ADHD), a highly heritable neuropsychiatric disorder^147; 148^. Poelmans et al. found that the gene ontology process “calcium ion binding,” which plays an important role in neurite migration, was significantly enriched in the 14 ADHD-associated genes^149^. Genome-wide analyses have identified *MCTP1* single nucleotide polymorphism (SNP) in bipolar diseases^150^. There was also a report of *MCTP2* SNP in schizophrenia^151^.

Our gene based association analysis suggests a high proportion of causal variants in *MCTP2* exert effects in the same direction and indicates the association between variant burden and face recognition ability in the male cohort, at least in part, by the effects of *MCTP2*. It is likely that the MCTP2 protein functions in the CNS and it will be interesting to investigate whether its role in face recognition is a general feature in neural signaling or a specialized step in specific regions of the brain.

### Diagnosis of CP in Linkage Analysis

If both parents exhibit a disability in face recognition which could be caused by different genetic origins, it is more difficult to localize the causing genetic variations ^62; 65^.

For the families with clear inherited models, correct and consistent phenotypical diagnosis for each family member is very important before genetic analysis. Currently, while specific tests have proven useful ^152^, no single test is of sufficient discriminatory power for the entire spectrum of CP ^60; 83; 95; 152; 153^. It is difficult to assess the exact severity of the individual CP cases by behavioral tests, because behavioral adaption, other cognitive skills, experience with faces or testing forms balance the deficit to an unknown extent, even within a single family with similar genetic and environmental background. Mistaking a true CP as a normal one due to his/her normal performance in behavioral tests would undermine genetic studies.

In our behavioral tests for face recognition in family A, there was also a variable behavioral profile in individuals who experienced everyday difficulties in face recognition. For example, V:10, perfected normally on the CFMT-C (z = −0.06) and excellently on the FIE test (z = 2.90) (Table 1), but he was already aware of his own real-world face recognition difficulties while he attended college, before we contacted him.

Another representative example was VI:7, a distant relative of the index, whose performance was fairly good on the CFMT-C (z =1.26) and FIE (z = 1.48), and would not have been diagnosed as having CP if only according to the available behavioral tests. However, she depended on clues beyond the face when recognizing people (Table 1, more detailed information can be obtained by contacting the corresponding author). Neuroimaging study showed that the activation pattern in her rFFA was indeed defective both in the FIE (Figures S6g, 5a) and the adaptation paradigm (Figures 4g, 5b).

Considering the within and between familial variations among CPs ^61–64; 154^, and the fact that CPs must experience face recognition difficulties on a regular basis in typical situations where recognition is not problematic for individuals without face processing impairments, we adopted a strategy with two step linkage analysis: in linkage 1, we defined family members under 60 as cases of CP (CPs) with clear difficulties in face recognition revealed by the interview and performed poorly (−1.5 SDs) below the average level in at least one of the face behavioral tests; those over 60 not suitable for behavioral tests but with lifelong difficulties in face recognition also as CPs; to be cautious, those with normal performance on behavioral tests but obvious face recognition failure experience as unclear; and the remaining members as normal; in linkage 2, we added people who were set as unclear in linkage 1 but with real-world face recognition defects, also as CPs (Table S1). We detected the same region on chromosome 15q with LOD scores over 3 from both linkage analyses (Figures 1b, 1c, S1a, S1b).

### Mutations in the *MCTP2* Gene

The mutation encoding p.I716S we found in *MCTP2* in the linkage region on chromosome 15q fully co-transmitted over several generations with all 18 affected individuals in family A. This mutation is private to family A, not present in public databases or in our in-house data, albeit another rare variant rs200314451 (c.T2147C; p.I716T) has been reported at this location in public databases.

The p.I716S is located at the first TMR of MCTP2 protein (RefGene: NM_018349). The isoleucine at 716 is evolutionally conserved within primates (Figure S3b) which are highly developed in face processing ^10; 37–39; 155; 156^.

Results from the prototypical CP family A indicate that rare, and private, functional mutations in *MCTP2* could be related to the CP phenotype and prompted us to examine more CP individuals and families. This strategy is justified from knowledge of monogenic disorders, where it is typical for different rare variants with large effects of the same gene to segregate with disease in different families. We screened 75 individuals with the most serious self-reported face recognition abnormalities at one end of the population distribution through the questionnaire from a cohort of 2904, rather than simply verifying them among the people who actively reported their deficient face recognition. Five rare heterozygous variants including one frameshift and four missense mutations were found in seven individuals (Figures 2a, S3a, Table S8), Their difficulties in daily face recognition were confirmed by detailed interviews. All the available family members also showed a clear phenotype-genotype correlation to exclude the observation of *MCTP2* variants by chance. This suggests the significance of rare functional mutations in *MCTP2* in CP. The association of the alleles in *MCTP2* that harbor rare coding variants of moderate or large effect on the protein coding with face recognition ability was further supported in males by a gene based burden analysis from a second cohort of 1928 individuals (Table 2). These results suggest a high proportion of causal variants in *MCTP2* exert effects in the same direction in the male cohort, beyond chance.

The frameshift deletion c.239delG, p.S80fs, in *MCTP2* which was found in three CPs from the 75 top candidate CPs, is likely to eliminate the MCTP2 protein. In our second cohort, the ExAC and the gnomAD databases, the MAFs in the population of East Asian for this mutation were 0.0026, 0.0031 and 0.0045 respectively, which lies within the reported range of CP prevalence and is high enough to attempt the identification of additional carriers consenting to a characterization of their face recognition behavior through a reverse-phenotyping approach. We detected 16 individuals carrying this mutation in a heterozygous condition with a MAF of 0.0046 in a third independent cohort of 1757. Among them, differences in the behavior of daily face recognition were detected between 14 carriers and 19 non-carrying volunteers (Table 3). 4 were clearly aware of their difficulties in face recognition before we contacted them, one felt different from others in recognizing faces, 8 individuals developed adaptive behaviors and used explicit learning strategies for person recognition, only one did not show obvious abnormal face recognition ability during the whole interview (Table S11). The individual differences in phenotype outcomes may be modulated both on the neurodevelopmental levels and environmental factors with other possible interactions such us more genetic traits, cognitive skills and learned compensatory strategies. Family members with the p.S80fs mutation further confirmed the phenotype-genotype correlation (Table S11, more detailed diagnostic information can be obtained by contacting the corresponding author).

### fMRI Analysis

fMRI is helpful both as a diagnostic means for endophenotype and for mechanistic link between genotype and brain activity.

We have focused on the rFFA because of its involvement both in detecting the presence of a face and in discriminating different faces ^33; 34; 99–104^.

In the fMRI analysis, the activation of rFFA in Families A and 3-2 without mutations in *MCTP2* was similar to that of the control group, in face detection, upright face recognition, inverted face recognition, different or the same individual face identification. The rFFA of members with the I716S in family A or the S80fs in family 3-2 also showed normal activation during face detection, suggesting these *MCTP2* mutations did not affect face detection. However, in the fMRI adaptation paradigm based on the theory that neural responses to faces are attenuated across repeated presentations of the same stimuli, an effect known as repetition suppression ^157–159^, the activation of the rFFA in all mutation carriers to paired identical faces was not lower than that of paired different faces, compared with that of normal people or non-carriers in families. These results indicate, that the mutations in *MCTP2* caused a failure in repetition suppression in the rFFA as well as behavioral difficulties in identifying previously encountered faces. In addition, the rFFA of A-VI:7 and 3-2-III:4 showed a significantly different activation pattern during the presentation of upright faces and inverted faces for the FIE. The rFFA of A-VI:7-was less active to the upright faces than the inverted faces, while the rFFA of 3-2-III:4 was less active to both types of faces.

Three fMRI subjects with the I716S mutation in family A and the one with the S80fs in family 3-2 had difficulties in recognizing people in daily life. In the fMRI study, when they were repeatedly presented with the same face, the repetition-suppression phenomenon in the rFFA was not obvious. It will be helpful if more families with *MCTP2* mutations are examined by fMRI.

We need to further investigate whether CP patients with *MCTP2* mutations have problems in specific adaptive mechanism of facial processing, or problems in other visual stimulation processing in general. Although in some non-face behavioral tests, these CPs performed normally, such as the CCMT test.

Our genetic results provide evidence that *MCTP2* gene mutations underlie CP and our fMRI results suggest that *MCTP2* is involved in neural circuits required for distinguishing different faces.

## SUBJECT DETAILS

### Participants

This study has been approved by the Committee for Protecting Human and Animal Subjects at Peking University (#2015-06-01e). Written informed consent to participate in the research study and to have the results of this research work published was obtained from participants or their legal representatives prior to any tests. Genomic DNA was isolated from whole blood (BD Vacutainer 3.2% Sodium Citrate (1:9)) using the Gentra Puregene Blood Kit (QIAGEN) or saliva using the GeneFiX DNA Saliva Collector and Isolation Kit.

### Family A

The proband V:11in family A was ascertained by self-referring. Initially, 35 available family individuals (18 CPs, including 9 males and 9 females, aged from 16 to 72; 17 non-CPs, including 9 males and 8 females, aged from 15 to 78) were assessed based on a standardized semi-structured interview ^54; 60; 160^. 25 family members younger than 60 years of age performed the CFMT-C ^80^, the matched CCMT ^81^ and the FIE Discrimination Test ^82^. 34 individuals (not including VI:9) were included in the linkage study. Four individuals (V:6, V:9, V:11, and V:19) were selected for the WES. Two individuals (V:19 and VI:5) were selected for the WGS. This information is listed in Table S1.

### Additional CP Individuals with Mutations in *MCTP2*

To look for more CP individuals and examine whether rare functional mutations in *MCTP2* are associated with CP, 2904 individuals (average age of 19.25 ± 1.30, 2161 females, 743 males) from the Westlake BioBank for Chinese (WBBC) pilot project ^161^, were screened with a questionnaire adapted from the 20-item self-report measure for quantifying CP traits. 78 individuals scored worse than the mean by 2 SDs. 75 of them provided DNA samples. 44 individuals including 7 individuals with *MCTP2* mutations agreed to an interview for CP diagnosis and family members were contacted for availability of further studies.

### Samples for Gene Based Association Analysis

The second cohort of 1928 individuals (average age of 18.51 ± 0.93, 1085 females, 843 males) used in the association of the rare functional alleles in *MCTP2* with face recognition ability were also from the WBBC project ^161^. The coding sequences of the *MCTP2* gene for each person was analyzed by tagged-amplicon deep sequencing.

### Individuals with the c.239delG, p.S80fs Mutation in *MCTP2*

A third cohort of 1757 individuals (average age of 19.13 ± 1.07, 1295 females, 462 males) were sequenced for the presence of the c.239delG, p.S80fs mutation in the exons of *MCTP2*. Individuals and family members carrying this mutation were further contacted and assessed by the standardized semi-structured interview. The coding sequences of the *MCTP2* gene for each person was analyzed by tagged-amplicon deep sequencing. The differences in the behavior of daily face recognition between the 14 carriers and 19 non-carrying volunteers from the same cohort were analyzed by independent sample t-test.

### Control Samples for Behavioral Tests

338 normal participants (average age of 42.68 ± 16.53, 164 females, 174 males) were tested. These participants were unrelated. Participants were not selected for face recognition ability except for known history of major brain injury, or other major disorders likely to affect face recognition (e.g., Alzheimer’s disease), representing a random sample of the community. They were tested on a battery of tests including the CFMT-C, the CCMT, the FIE Test and the Cambridge Face Perception Test -Chinese (CFPT-C).

### fMRI Subjects

Age-related dedifferentiation and compensatory changes in the functional network underlying face processing have been found in studies ^154; 162–167^. Therefore, in order to exclude age influence on fMRI results, we mainly studied family members younger than 30 years old. Family members who participated in the fMRI experiment were on a voluntary basis. In family A, 3 CP individuals (VI:5, VI:7 and VI:9) and 3 non-CP individuals (VI:1, VI:2 and VI:10) took part in fMRI; in family 3-2, III:4 (CP) and III:2 (non-CP) took part in fMRI.

21 non-CP students (average age = 23.63 ± 3.71, 6 females, 15 males) were recruited for fMRI analysis.

CP referred to those with obvious face recognition problems in daily life diagnosed by the structured interview.

All participants reported normal or corrected to normal vision, no history of neurological or psychiatric conditions and all were right handed. Anatomical volumes (i.e., structural MRIs) had been routinely checked. One of the normal students was excluded from neuroimaging studies because the maximum head rotation was over 1.5 degree or the maximum translation was over 2 mm during localization.

We adopted the methods of studying AP (acquired prosopagnosia) cases that the severity of sporadic cases could be reported by comparison with the control population ^138; 168–172^. In fMRI analysis, each family member was compared individually to a small sample of normal controls by a modified t-test (Crawford and Garthwaite, 2002; Crawford and Howell, 1998).

## METHODS DETAILS

### Structured Interview for CP Diagnosis

The diagnosis of CP was based on a standardized semi-structured interview (Table 1, S7 and S10) ^54; 60^, which documented a variety of impressive qualitative differences between CP and non-CP, and had been validated with objective face recognition tests in previous studies ^60; 160^. These criteria were employed in recent literature on CP ^69–79^.

The interviewer asked questions in a semi-structured interview format with three or four questions about each diagnostic item. Interviews were held to embed the questions into conversations and questions about the same diagnostic items not asked sequentially. Interviews included a medical history in order to exclude conditions which might cause or mimic prosopagnosia. Affected people present a lack of confidence with face recognition. Symptoms include lasting and irritating subjective uncertainties of face recognition, failure to recognize familiar people out of context or in crowded places, overlooking familiar people and confusing strangers with familiar people, face recognition/learning time longer than socially accepted, development of adaptive behavior of critical situations and strategies for visual recognition of people and time of onset.

Consistent with the interview results, some individuals were aware of their CP before we contacted them. Some individuals who could identify people via non-facial features such as voice, gait and general appearance and manner, or were unaware of face recognition problems, but had developed obvious compensatory strategies to cope with difficulties were also diagnosed as CPs.

### CP Questionnaire

To effectively screen CP candidates from big samples, we adapted a Chinese 20-item version from the 20-item self-report measure for quantifying prosopagnosic traits ^95^, which asks about tangible experiences. This 20-item Questionnaire was included in a set of questionnaires for many research purposes and filled out by two cohorts of individuals online. Invalid questionnaires were dropped due to no distinction between forward and reverse questions. 2904 valid questionnaires for the first cohort and 1928 valid questionnaires for the second cohort were collected. The internal reliability measured by Cronbach’s α was 0.828 and 0.902 respectively. In the first cohort, 343 individuals finished the questionnaire for a second time several weeks later and the correlation for the first and second results across each individual was very high by Pearson correlation coefficient analysis, r = 0.081, *p* < 0.001. There was a significant difference of the score distribution between the female and male participants (*p* < 0.001, two tailed t test), so the candidate CPs were screened and the gene based association study were carried with different genders respectively. An individual with a score beyond 2 SDs of the mean of the controls was defined as a CP.

### Stimuli and Procedures of Behavioral Tests

All behavioral tests were adapted and integrated into a whole set using the Hyper Text Markup Language. All participants were tested individually. The tests were run on a desktop PC with screen resolution 1024 × 768, refresh rate 85 Hz. Participants were seated at a viewing distance of approximately 50 cm from the screen. All participants were tested wearing their usual optical correction. Participants were asked to confirm that they could focus without seeing blur on the computer screen. No participant reported any difficulty with focus at these distances. Gray-scale adult Chinese faces were used.

### Cambridge Face Memory Test-Chinese (CFMT-C)

The Chinese face version of CFMT ^80^ was kindly provided by Professor Jia Liu of Beijing Normal University which we integrated with other behavioral tests into a whole set, and performed according to the standard procedure including the practice phase, the “Learn” phase with 18 trials, the “Novel Images” phase with 30 trials and the final “Novel Images with Noise” with 24 trials. All faces were Chinese male, shown without hair or facial blemishes and with neutral expressions. Participants were instructed to press the key corresponding to the number 1, 2 and 3 below faces. The test includes a total of 72 trials. Scores were reported as percent correct across the full test.

### Cambridge Car Memory Test (CCMT)

CCMT is a test similar in the experimental design as CFMT, with stimuli replaced by whole cars ^81^. We used CCMT as a control of CFMT to test for potential general object recognition deficits and to quantify the individual ability of performing such kind of tests. The original version of CCMT was kindly provided by Professor Bradley Duchaine of Dartmouth College, USA, which we integrated with other behavioral tests into a whole set, and performed according to the standard procedure ^81^ including the practice phase, the “Learn” phase with 18 trials, the “Novel Images” phase with 30 trials and the final “Novel Images with Noise” with 24 trials. The stimuli were the modified computer-generated images of actual car makes and models from the original CCMT. Participants were instructed to press the key corresponding to the numbers 1, 2 and 3 below cars. The test included a total of 72 trials. Scores were reported as percent correct across the full test.

### Cambridge Face Perception Test-Chinese (CFPT-C)

We developed a version of CFPT using Chinese faces as stimuli (CFPT-C), according to the standard procedure ^65^ by morphing six different individuals with the target face, containing 88%, 76%, 64%, 52%, 40%, and 28% of the target face in turn. The Chinese faces were male, shown without hair or facial blemishes and with neutral expressions. All faces were photographs of Chinese students at Peking University, with written consent forms collected before photographing. Each individual was photographed in the same range of views and lighting conditions. Eight upright and eight inverted trials were intermixed, with the upright trial occurring first half the time. Participants had one minute to arrange six morphed faces according to their similarity to a target face by clicking on a face and indicating where that face should be moved by clicking in the area between two faces. Scores were computed according to the previous paper ^65^. The internal reliability measured by Cronbach’s α in our sample of 170 individuals was 0.705 for upright faces, and 0.483 for inverted faces. However, after our pilot study, the CFPT-C was excluded from further study, because strategies such as just comparing partial facial features were used in normal people as in CPs and inconsistent results were reported by others ^57; 65; 173^.

### Face Inversion Effect (FIE) Discrimination Test

The stimuli consisted of 20 gray-scale individual face images, cropped using the same 4X4.5cm oval window to remove cues from the hairline and face contour. All faces were photographs of Chinese students at Peking University, with written consent forms collected before photographing. Photographs were not repeatedly used in different tests. Pairs of face stimuli were presented sequentially either upright or inverted in a randomized order. The first face stimulus was presented in the upper-left quadrant of the screen for 0.5 s. After an interstimulus interval of 0.5 s, the second stimulus was presented in the lower-right quadrant for 0.5 s. The next trial will not begin until a same or different response was made by pressing one of two keys to respond by the participant. 80 trials were conducted in this test, half of them with upright faces and half with inverted faces. In each condition (upright or inverted), the chance with the two identical faces was 50%. A few practice trials were presented before the beginning of the experiment. Scores were reported as percent correct for each condition. For the measure of FIE, the difference in performance level between upright and inverted faces, an FIE index was calculated by entering the correction of performance for upright and for inverted faces in the following formula: FIE = (upright –inverted) / (upright + inverted).

### Data Analysis for Behavioral Tests

Due to possible effects of ageing and sex difference on the scores of CFMT-C, CCMT, upright faces and inverted faces in FIE and the FIE index, we used multiple stepwise regression analyses to identify the covariates (sex, age) specific to each trait from 338 participants. The results of CFMT-C and CCMT showed noticeable age-related decline and sex differences. The scores of upright faces and inverted faces in FIE correlated with ageing but not sex, while the FIE index was not affected by age or sex among the control samples.

Figure S7 shows a scatterplot of age against CFMT-C score (a) and CCMT score (b) including all participants (aged 15–83 years) in different sex. We fitted age-related curves to the CFMT-C and CCMT scores with different sex, as shown CFMT-C_female_ = −0.0087age^2^ + 0.4954age + 71.508,

CFMT-C_male_ = −0.0069age^2^ + 0.3159age + 69.506,

CCMT_female_ = −0.0073age^2^ + 0.5034age + 50.433,

CCMT_male_ = −0.0087age^2^ + 0.4954age + 71.508.

Figure S8 shows a scatterplot of age against upright face score (a) and inverted face score (b) in FIE including all participants (aged 15–83 years). We also fitted age-related curves to the upright face and inverted face scores, as shown Upright = −0.0051age^2^ + 0.2159age + 79.821,

Inverted = −0.0016age^2^ + 0.0173age + 63.813.

Examining function curves suggested that the behavior performance remained stable across early middle age, but began to decline noticeably at approximately 50 years of age. This is consistent with previous studies. In terms of the validity of the behavioral scores, only individuals with ages under 60 (average age = 36.51 ± 12.34, 130 females, 138 males) were included to calculate the best estimate of CP cut-offs.

Significant covariates for each behavioral score were considered for the adjustment of behavioral performance. We used the fit-and-residual procedure to calculate the standard residual of each participant (z score, a reflection of the behavioral performance) for CP family members as described previously ^57^. The regression function describing the relationship between age, sex and CFMT was: CFMT score = −0.212 age + 6.188 sex + 73.724. The difference between the participant’s CFMT score and the estimated score from the function given his/her age and sex (i.e. the residual) was calculated. The standard deviation of the controls’ residuals of CFMT score was: 11.189. Z score was calculated by dividing the participant’s residual by the standard deviation of the controls’ residuals.

The regression function describing the relationship between age, sex and CCMT was: CCMT score = −0.134 age −2.968 sex + 69.444. The difference between the participant’s CCMT score and the estimated score from the function given his/her age and sex (i.e. the residual) was calculated. The standard deviation of all the participants’ residuals for CCMT score was: 10.252. The z score was calculated by dividing the participant’s residual by the standard deviation of the controls’ residuals. The regression function describing the relationship between age and upright faces was: Upright score = −0.170 age + 87.641. The difference between the participant’s upright faces score and the estimated score from the function given his/her age (i.e. the residual) was calculated. The standard deviation of all the participants’ residuals for upright faces score was: 8.862. The z score was calculated by dividing the participant’s residual by the standard deviation of the controls’ residuals. Participants with z score below 2 SDs were considered performing poorly in these tests.

For the FIE index, there was no correlation with age or sex. Normalized z score was calculated for each subject by subtracting the mean of the control sample and dividing by the control samples’ SD, and was considered as performing poorly if below 2 SDs.

## GENETIC ANALYSES

### Genome-wide Genotyping for Linkage Analysis

DNA samples were genotyped using Infinium Human OmniZhongHua BeadChips (Illumina), and normalized bead intensity data obtained for each sample were converted into SNP genotypes using Genome Studio. SNPs were then selected according to the following parameters: genotyping rate > 95%, minor allelic frequency > 1% and no significant deviation from Hardy–Weinberg proportions (*p* > 0.001) by PLINK software ^174^. Gender corresponding to each DNA sample was checked by analysis of X chromosome heterozygosity using PLINK. The initial Mendelian inheritance in family A0001 was analyzed by PLINK and KING toolset175.

### Data Analysis for Linkage Study

Parametric linkage analysis was performed with the Merlin programs ^176^, assuming autosomal dominant inheritance with 80% penetrance (for linkage 1, 90% for linkage 2), disease allele frequency 0.001, and phenocopy rate 0.05. Because the presence of linkage disequilibrium (LD) might inflate linkage statistics, LD maps were constructed with the PLINK tool within family members with the LD thresholds (r^2^ < 0.5). Following data QC, 177,126 (r^2^ < 0.5) informative SNPs were selected for linkage analysis with Merlin ^176^. Because the Merlin software could analyze a large set of polymorphic markers in a limited number of family members, family A was split into 2 sub-families with common ancestors. 1000 simulation analyses were performed to test for the statistical power of the significant results under the null hypothesis of no linkage, with simulated data, while maintaining the pedigree structure, allele frequencies, and recombination fraction. By convention, parametric linkage results were reported in LOD scores. A LOD score of > 2.5 was taken as being highly suggestive of linkage while a LOD score of > 3.0 was taken as convincing evidence of linkage. Haplotype construction was performed using the SimWalk2 program ^177^.

### Generation of CNV Calls

Genotyping and signal intensity data (genotype, GType; Log R ratio, LRR and B allele frequency, BAF) were exported from the GenomeStudio software 2011.1 (San Diego, CA, USA). The subsequent CNV calling analyses were performed using PennCNV (v.2011 Jun16) ^178^ according to the manual. Minimum number of markers of 3 were set as threshold for all CNV calling. The samples met the following QC measures were thought with good quality for further analysis: (1) within the standard deviation (SD) of the normalized intensity (LRR < 0.35); (2) |GC bp wave factor (GCWF)| < 0.05; (3) with CNV call count < 70.

### Whole Exome Sequencing (WES)

WES was performed by the Next-Generation Sequencing Center of Biomedical Pioneering Innovation Center, PKU. Exome capture was performed using Agilent SureSelect Human All Exon V4 capture kit. 100 bp paired-end sequencing was done on Illumina HiSeq 2500.

### Data Analysis for WES

Sequences were aligned to human genome build hg19 using BWA ^179^. Duplicate reads were marked using Picard (http://broadinstitute.github.io/picard/). Genome Analysis Toolkit (GATK) ^180–182^ was used for base quality score recalibration (BQSR) and local realignment around indels to refine alignment artifacts around putative insertions or deletions. Variant discovery was performed in two steps: variant calling with GATK HaplotypeCaller followed by joint genotyping using GATK GenotypeGVCFs. The resulting variant call set was refined using Variant Quality Score Recalibration (VQSR) as implemented in GATK VariantRecalibrator according to the GATK Best Practices recommendations ^181; 182^. The VQSR scores were used to define low quality variants for downstream processing. If the coverage in MCR was under 20-fold depth in all the 4 CP individuals investigated by WES, we performed direct Sanger sequencing to verify these regions. All variants were annotated to RefSeq hg19 and ANNOVAR ^183^ were used to add alternative allele frequencies, variant effect predictions and functional annotations.

### Whole Genome Sequencing (WGS)

WGS was also performed by the Next-Generation Sequencing Center of Biomedical Pioneering Innovation Center, PKU. Sequencing libraries were built with NEBNext Ultra DNA Library Prep Kit for Illumina. 150 bp paired-end sequencing was done on Illumina HiSeq 4000.

### Data Analysis for WGS

FastQC (http://www.bioinformatics.babraham.ac.uk/projects/fastqc/) was applied to perform quality checks on various aspects of sequencing quality. Low quality bases were removed by Trimmomatic software ^184^ with the parameters HEADCROP:5, LEADING:25, TRAILING:25, SLIDINGWINDOW:4:15 MINLEN:35. After that, clean sequences were aligned to human genome build hg19 using BWA ^179^. Duplicate reads were marked using Picard (http://broadinstitute.github.io/picard/). SNPs and INDELs were called by GATK HaplotypeCaller ^180^. DELLY ^185^ and BreakDancer (http://www.nature.com/nmeth/journal/v6/n9/abs/nmeth.1363.html) were used to discover structural variants. BAM files locally realigned around INDELs were used to determine the average coverage, using GATK DepthOfCoverage package and default settings. If the coverage in MCR was under 20-fold depth in all the 2 CP individuals investigated by WGS, we performed direct Sanger sequencing to verify these regions. All variants were annotated to RefSeq hg19 and ANNOVAR^183^ were used to add alternative allele frequencies, variant effect predictions and functional annotations.

### Tagged-Amplicon Deep Sequencing

Target-specific primers for the coding sequences of the *MCTP2* gene (NM_018349) were designed with universal primer sequences (termed CS1 and CS2) appended at the 5’-end and sequencing was performed as described previously ^186^.

### Data Analysis for Tagged-Amplicon Deep Sequencing

The raw paired 150 bp-long reads were mapped to the human reference genome (build hg19) using BWA. GATK was then used to perform local realignment and recalibrate base quality scores, producing a BAM file for each individual. The raw single nucleotide variants (SNVs) were filtered based on the variant quality score recalibration module in GATK. All variants were annotated to RefSeq hg19 and ANNOVAR^183^ were used to add alternative allele frequencies, variant effect predictions and functional annotations. We manually inspected each mutation using the Integrative Genomics Viewer ^187; 188^ to rule out false positive findings. If the coverage for each exon was under 500-fold depth in each individual, we performed direct Sanger sequencing to verify these regions.

### Sequence Validation

Mutations were amplified by PCR and validated by direct Sanger DNA sequencing. All reactions were 100% successfully validated. Primer sequences for PCR amplification and Sanger sequencing are included in Tables S5, S7.

### Founder Origin Testing

We genotyped nine multi-allelic microsatellite markers (CHLC.ATA22D04, AFMB077YD5, GATA128A02, AFM072YB11, AFM357TD9/D15S1038, CHLC.GATA73F01, GATA161C02, AFM217ZG1, AFM309VG9) around the c.239delG (p.S80fs) mutation of *MCTP2* from 94255659 bp to 96210769 bp on chromosome 15 in families 1-1, 3-6, 3-2, 3-8, 3-5 and 3-3. The inferred haplotypes were estimated by comparison to these microsatellites data collected on mother-father-offspring trios to test the possibility of the same founder origin.

## NEUROIMAGING

### Stimuli

In the localizer experiment, images of faces, non-face objects (e.g., chairs, food, and tools), and texture patterns (scrambled faces) were presented at the center of the screen, subtended 6.2°×6.2°. In the adaptation paradigm, the stimuli were gray-scale images of young Chinese men (hair cropped with neutral expressions), the same as those used in the FIE behavioral test. The stimuli were presented by a MRI compatible projector system (SA-9900, The Shenzhen Sinorad Medical Electronics Co., Ltd, China, http://www.sinorad.com), with a spatial resolution of 1,024×768 and a refresh rate of 60 Hz.

### Experimental Design and Procedures

Each participant completed the same single scan session consisting of one functional localizer run and five runs for the adaptation paradigm. During the experiment, participants lay on their back in the scanner, using earplugs to reduce noise and sponges to hold their heads in place to reduce head movement. Participants viewed the stimuli presented on a translucent screen visible via a mirror mounted to the head coil at a distance of 60 cm. In the localizer run which lasted 360 seconds (s), with a 12 s dummy at the beginning of the run, images appeared at a rate of 2 Hz in blocks of 12 s, interleaved with 12 s blank blocks. There were five blocks for each type of images in the run. Each image was presented for 300 microseconds (ms), followed by a 200 ms blank interval. Subjects performed a one-back task during scanning to ensure maintenance of attention to the stimuli. In the adaptation paradigm, pairs of face or house stimuli were presented sequentially either in the upright or the inverted manner in a randomized order that was optimized for the extraction of the hemodynamic response in an event-related fast presentation design. Each trial lasted 2000 ms. The first and second stimuli were presented for 250 ms, with an interval of 500 ms and followed by 1 s fixation. Blank screen with white cross fixation point was set between trials, with a presentation time of a random even number in the range of 0-10 s to optimize the efficiency of the event-related fMRI design by optseq2 (http://surfer.nmr.mgh.harvard.edu/optseq). Each run lasted 332 s, containing eight conditions (upright same face, upright different face, inverted same face, inverted different face, upright same house, upright different house, inverted same house and inverted different house). Five runs were included, each containing 12 trials for each condition. Subjects made a same/different response on each trial. The purpose we used an event-related design instead of block design of the adaptation paradigm, was to mimic the FIE behavioral test used to explore the ability of face recognition in CPs based on computers. Here we focus only on the face stimuli.

### fMRI Scanning

The fMRI data were collected in a 3T GE MR 750 scanner, with an 8-channel phase-array head coil (GE Healthcare, Waukesha, WI) at Peking University Center for MRI Research.

The gradient-echo echo planar imaging (EPI) sequence was employed for BOLD images acquisition, and the imaging parameters were as below: repetition time (TR) = 2000 ms, echo time (TE) = 30 ms, field of view (FOV) = 224mm × 224mm, matrix = 64 × 64, flip angle = 90°, slice thickness = 3.5 mm with 0.7 mm spacing, voxel size = 3.5 × 3.5 × (3.5 + 0.7) mm, 33 oblique slices covering the whole brain).

The structure images were acquired by a 3D inversion recovery-prepped T1-weighted sequence (fSPGR, sagittal acquisition, TR = 6.65 ms, TE = 2.92 ms, TI = 450 ms, flip angle = 12°, FOV = 256 mm × 256 mm, matrix = 256 × 256, 192 continue slices with 1mm slice thickness, voxel size = 1 mm × 1 mm × 1 mm).

### Data Analysis for Neuroimaging Studies

Preprocessing and data analysis were performed with SPM12 (Wellcome Trust Centre for Neuroimaging, London; http://www.fil.ion.ucl.ac.uk/spm/software/spm12/). Functional images were sequentially processed in accordance with the standard SPM approach as follows: interpolated to correct for slice timing, realigned to the middle volume, co-registered to structural scans using the mean functional image, spatially normalized to a standard echo-planar image (EPI) template based on the Montreal Neurological Institute reference brain template (MNI152, Asia brain), and spatially smoothed with an isotropic 8 mm full width at half-maximum Gaussian kernel. For anatomical reference, the statistical maps computed were overlaid to the 3D T1-weighted scans. First, face-selective regions were localized in each subject by BOLD signals. By comparing the “faces” condition with the “non-face” condition in the localizer experiment at the first-level analysis, we assessed this face-selective ROI for every participant by building a 5-mm radius sphere surrounding the coordinate of the maximum activation in rFFA at a threshold *p* < 0.05, the family-wise error (FWE) correcting for multiple testing, with MARSBAR followed by visual confirmation of their anatomical location. For participants who lacked a face-selective area with this criterion, we explored liberal uncorrected significance thresholds (as liberal as *p* < 0.05), to avoid missing effects that might be apparent at less stringent uncorrected thresholds. Because faces are processed more dominantly in the right hemisphere and because the ROIs in this study were localized more consistently in the right hemisphere, we chose to restrict our ROI-based analyses to regions in the right hemisphere (unless noted otherwise). Second, the above-defined ROI were tested for FIE using the contrast (upright different face > inverted different face) and adaptation to facial identity using the contrast (up different faces > up identical faces) by BOLD signals at the first-level analysis. Third, the time course of percent signal change from baseline fixation was extracted from the ROI for each condition in each individual with MARSBAR, and was plotted for each condition in controls (±SD) and family members by an in-house Matlab programme. Fourth, in order to directly compare each CP or non-CP with the control subjects, the percent signal change in the ROI was computed for each condition. Three data points around the peak of the hemodynamic response defined individually were averaged to estimate the percent signal change. Fifth, the percent signal change was used to compute both the FIE (upright different face – inverted different face) and adaptation scores (upright different face – upright identical face) for each subject, allowing a comparison between both the CP and non-CP family members with the control group in each experiment by means of Z-scores and the modified t-test score ^105; 106^.

Results were visualized using xjView toolbox (http://www.alivelearn.net/xjview) in addition to built-in visualization in SPM12.

## Quantification and Statistical Analysis

Where applicable, statistical parameters including sample size, precision measures (standard error or standard deviation) and statistical significance are reported in the figures and corresponding legends. *P*-values of less than 0.05 are significant.

## Data Availability

Further information and requests for raw data, analysis details, and DNA samples may be directed to and fulfilled in compliance with IRB protocol requirements by Yi Rao. Whole exome/genome sequencing data are not deposited in a public repository because the genetic sharing plans of the IRB approved consent forms used in this study do not permit the deposition of such data into public or controlled-access databases.

## ACKNOWLEDGEMENTS

We are grateful to participants of this project (especially the proband and members of A0001 family), to Dr. Ingo Kennerknecht at the University of Muenster, Germany for sharing the interview and for helpful discussions, to Dr. Jia Liu of Beijing Normal University for sharing the Cambridge Face Memory Test-Chinese version and helpful discussions, to Dr. Bradley Duchaine of Dartmoutth College for sharing the Cambridge Car Memory Test and helpful discussions, to Dr. Jurg Ott, Shihui Han and Taoyu Wu for helpful discussions, to Dr. Pianpian Zhao of Westlake University, Drs. Jing Jiang and Kunjun Mao of Shangrao Medical College, Drs. Liang Chen and Jieyu Chen of Southern Medical University for help with sample collection, to Drs. Zi-Bing Jin and Xue Zhang for critical comments on the manuscript, to the High Performance Computing Platform of Peking/Tsinghua Center for Life Sciences (CLS) for computer support, to the CIBR, the CLS, the Shenzhen Bay Laboratory and the National Natural Science Foundation of China for funding support.

## AUTHOR CONTRIBUTION

YR initiated the idea of genetic analysis of higher cognition in humans, using CP as an example and supervised the project. YR and YS designed the project and wrote the manuscript. YS and WWM carried out fMRI. HFZ, WF and YS collected and coordinated the collection of samples. YS and WF performed CP diagnosis. YS, WF and EXZ did behavioral tests and data analysis. EXZ wrote software programs for behavioral tests. YS and ZQL performed linkage analysis. YS and WY carried out sequencing analysis. HFX was our collaborator in the Westlake BioBank for Chinese (WBBC) pilot project. YS performed founder origin analysis.

## Supplementary Information

**Figure S1:**
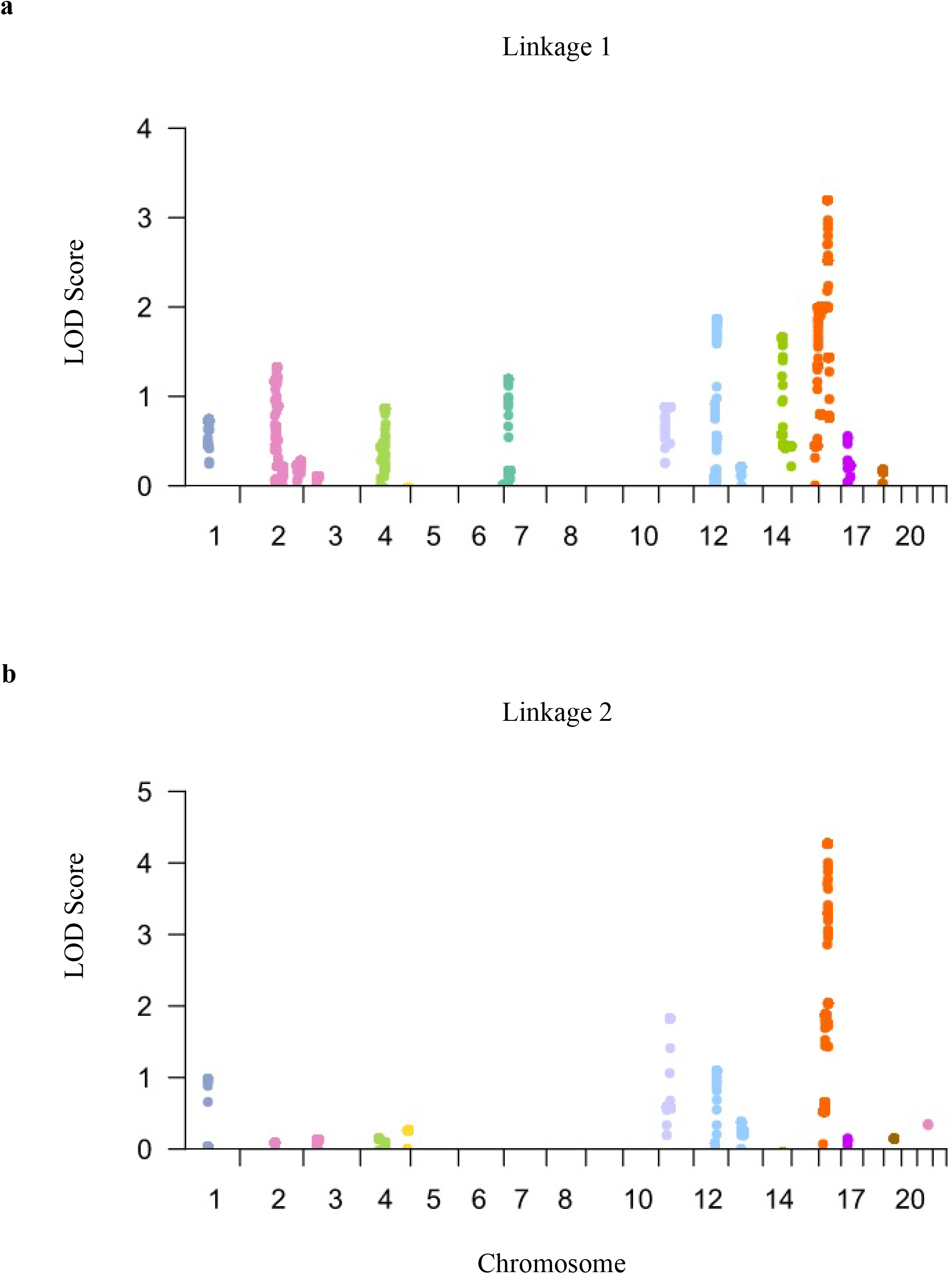
Genome-wide Screen Results of CP-susceptibility Loci. Allele-sharing multipoint LOD scores are shown. (a) for linkage 1, and (b) for linkage 2.

**Figure S2:**
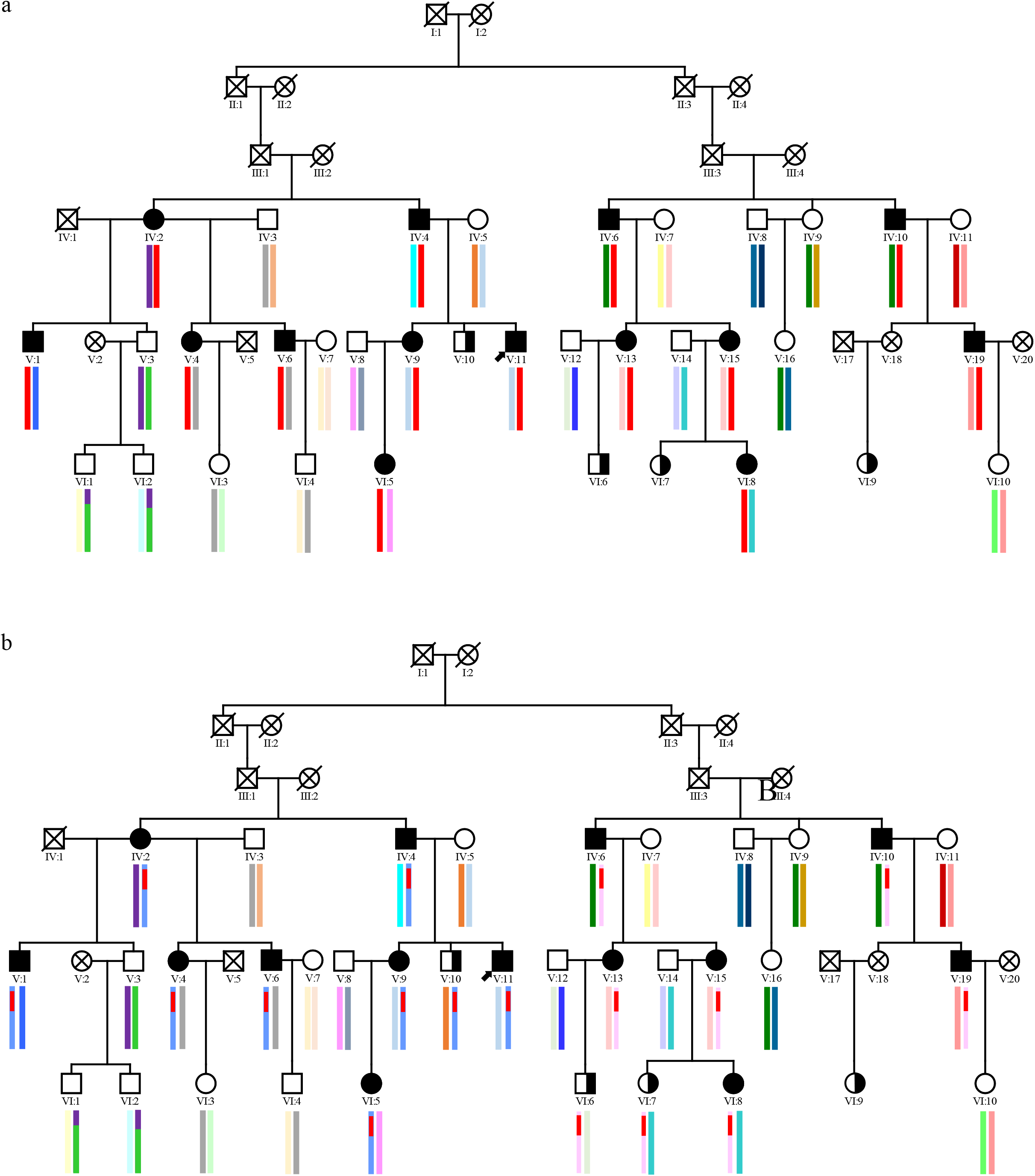
Haplotype Analysis for MCR on Chromosome 15q. (a) 315 markers with LOD scores over 3 in linkage 1 were used in haplotype analysis. Haplotypes are color-coded, with red indicating co-segregating with CP. (b) 400 markers with LOD scores over 3 in linkage 2 were used in haplotype analysis. Haplotypes are color-coded, with red indicating co-segregating with CP.

**Figure S3:**
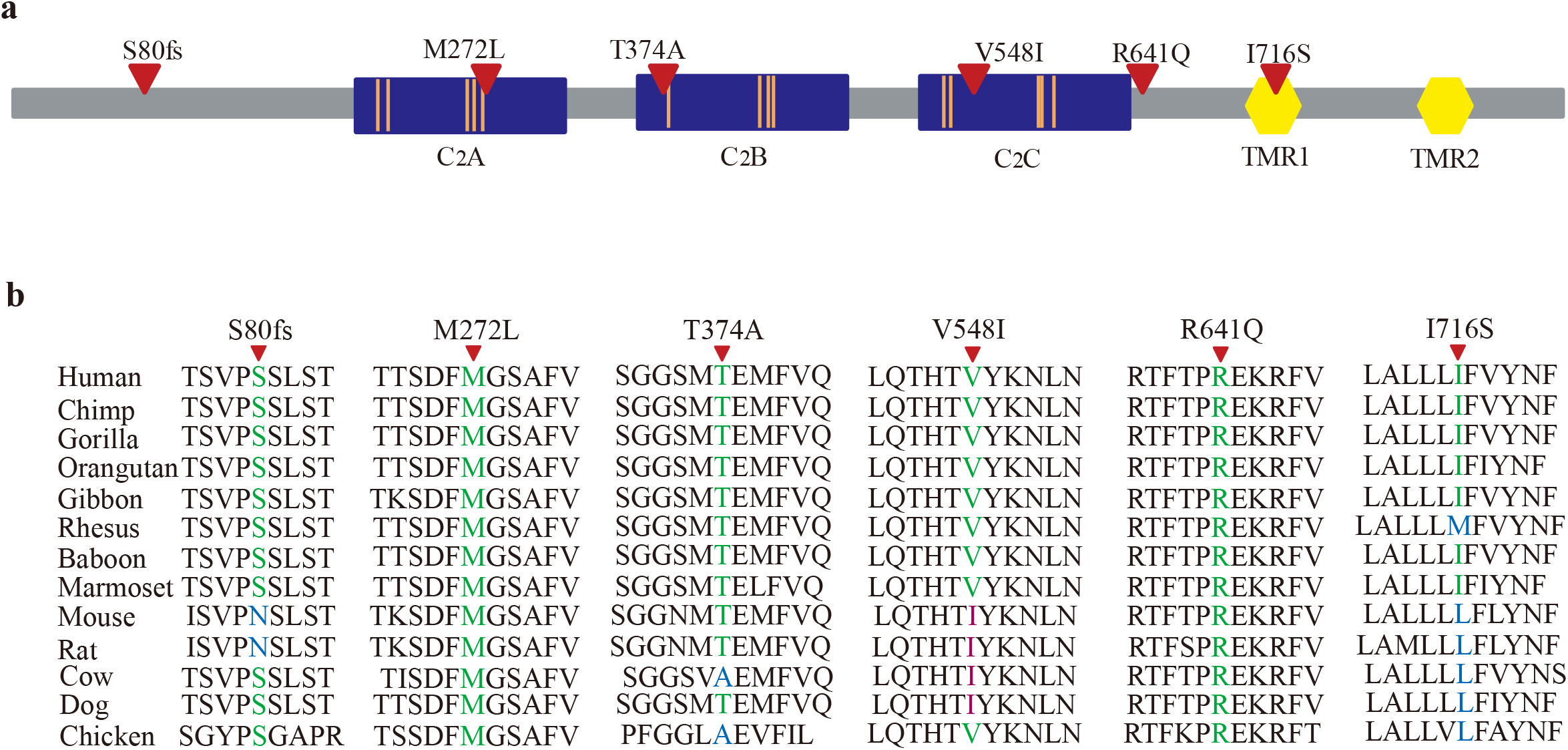
The Positions in the MCTP2 Protein Corresponding to MCTP2 Mutations in CPs. (a) The domain plot of the MCTP2 protein and locations of alterations found in CPs. The blue rectangles represent C2 domains, the yellow hexagons represent transmembrane segments; the orange lines indicate putative Ca2+ binding sites; the red triangles indicate amino acid changes. (b) Evolutionary conservation of amino acid residues in the MCTP2 protein in different species. Amino acids corresponding to mutations found in CP subjects are denoted in different colors. Green: identical to those in the human MCTP2, blue: non-identical to those in human MCTP2, fuchsia: non-identical amino acids but the same as the mutated in human MCTP2. Mutated amino acid residues are indicated by triangles on the top.

**Figure S4:**
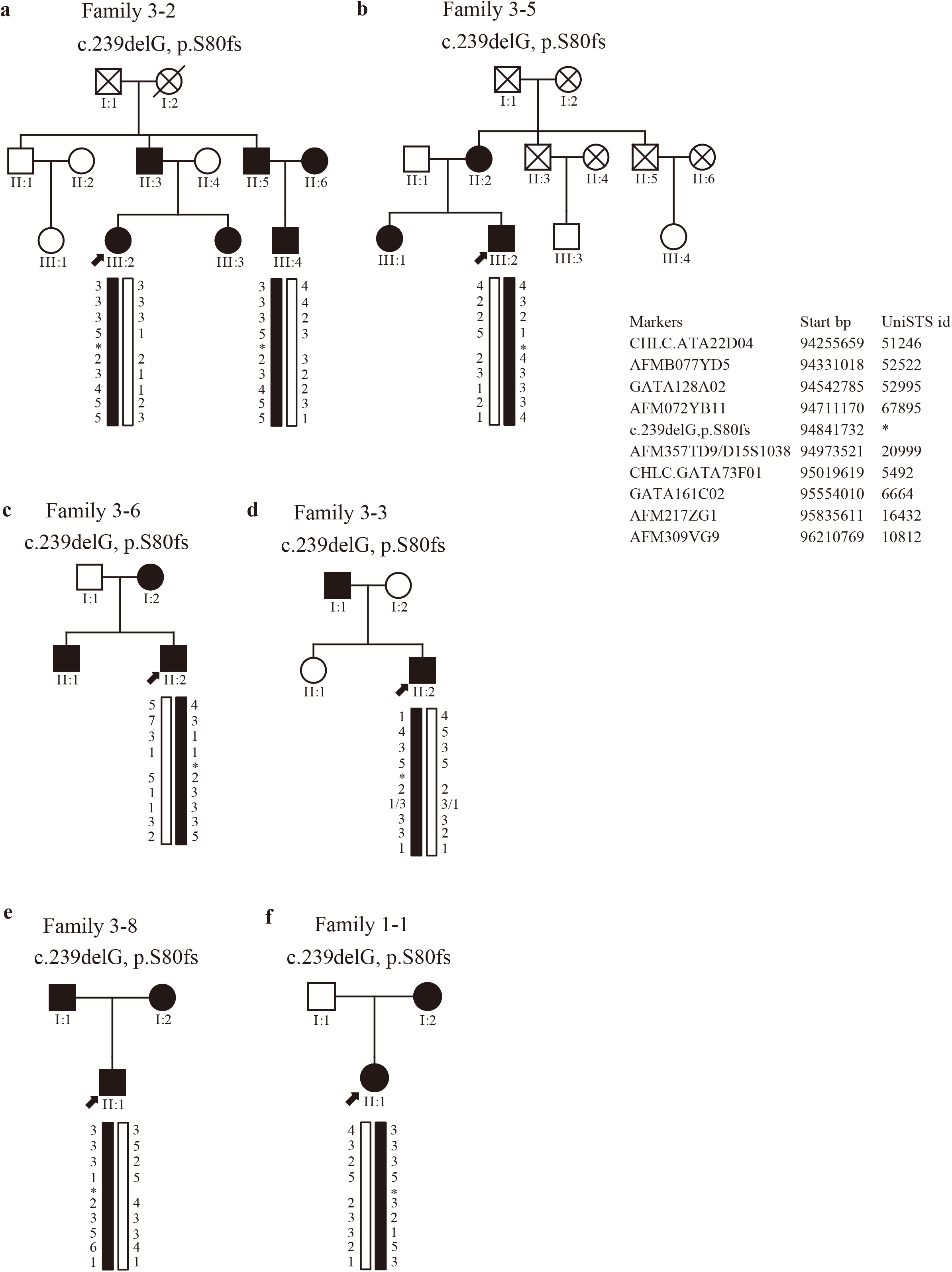
Inferred Haplotypes around the c.239delG (p.S80fs) Mutation. These were estimated by comparison to microsatellites data collected on mother-father-offspring trios. Haplotypes coded in black contain the c.239delG (p.S80fs) mutation. The haplotype on the left is paternal and the haplotype on the right is maternal.

**Figure S5:**
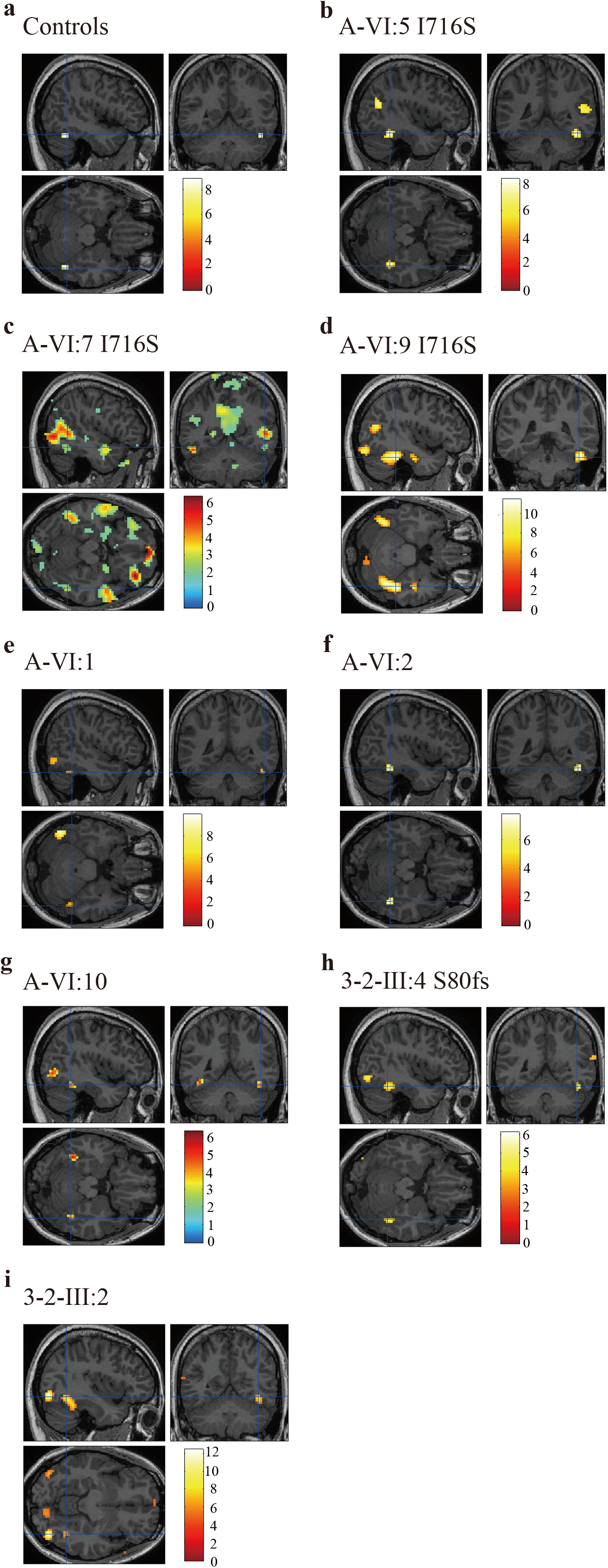
Brain fMRI from the Localization Experiment: Identifying the FFA ROI. Statistical parametric maps by contrasting the response to faces versus non-face stimuli, showing significant face-specific bilateral or unilateral activity in the FFA. Color scale represents statistical values of a one level t test comparing the fMRI signal while subjects viewed blocks of faces versus blocks of non-face stimuli. (a) rFFA responses in normal controls, pFWE < 0.05. (b) rFFA responses in A-VI:5 CP member with the I716S mutation, pFWE < 0.05. (c) rFFA responses in A-VI:7 CP member with the I716S mutation, p < 0.05. (d) rFFA responses in A-VI:9 CP member with the I716S mutation, pFWE < 0.05. (e) rFFA responses in A-VI:1 non-CP member, pFWE < 0.05. (f) rFFA responses in A-VI:2 non-CP member, pFWE < 0.05. (g) rFFA responses in A-VI:10 non-CP member, p < 0.001. (h) rFFA responses in 3-2-III:4 S80fs CP member, p < 0.0001. (g) rFFA responses in 3-2-III:2 non-CP member, pFWE < 0.05.

**Figure S6:**
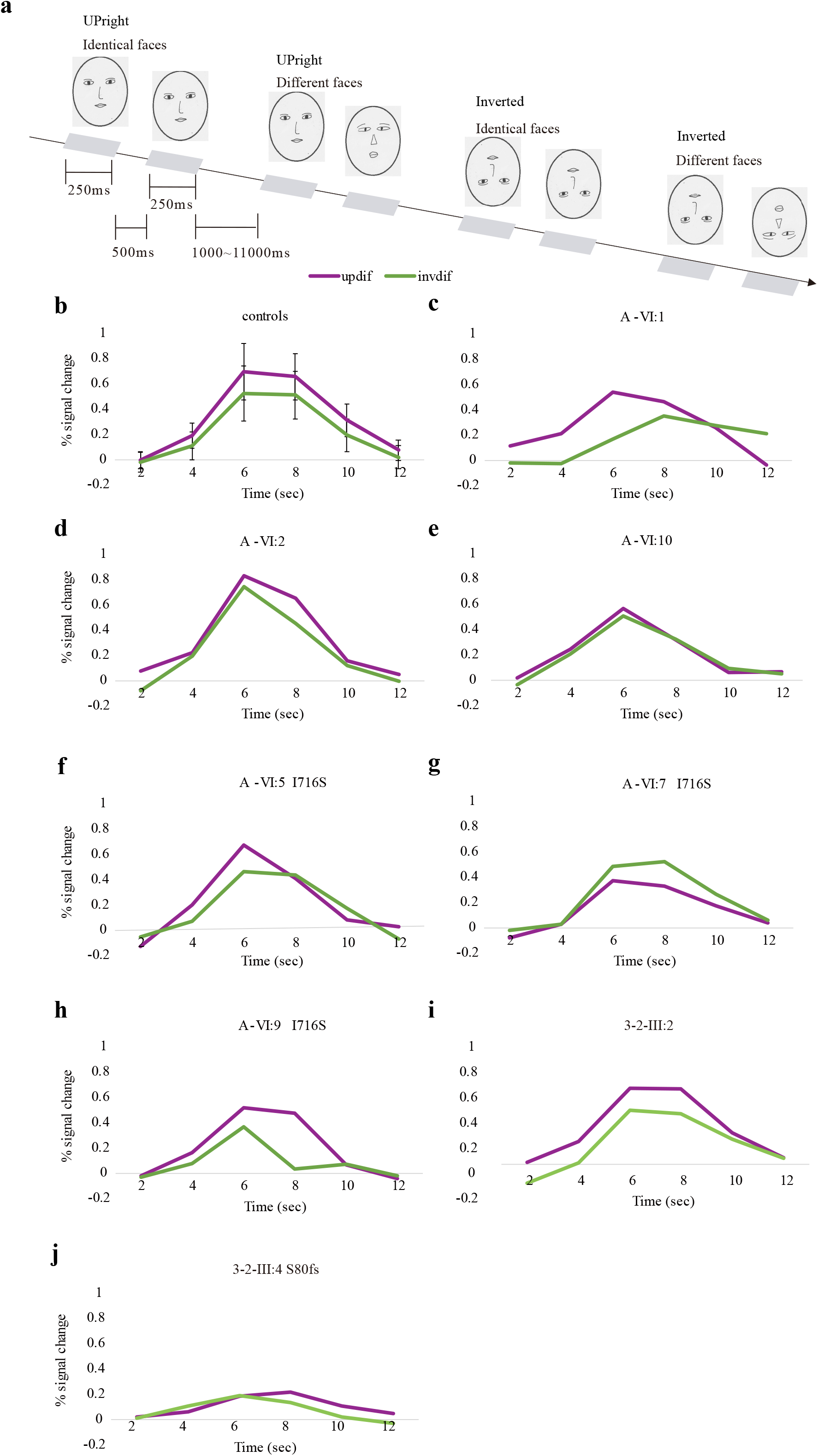
rFFA Activation in Face Sensitivity. (a) Experimental paradigm. Gray-scale photographs of 24 pairs of upright or inverted faces were presented sequentially in a random order during the experiment. Cartoon faces are used as examples. The average percent signal change (±SD for controls) from baseline fixation is plotted for the upright and the inverted face conditions. Bold response to pairs of upright different faces or inverted different faces in the rFFA of controls (n=20) (b); non-CP members A-VI:1 (c), A-VI:2 (d), A-VI:10 (e) and CP members from family A with the I716S mutation in MCTP2: A-VI:5 (f), A-VI:7 (g), and A-VI:9 (h); non-CP 3-2-III:2 (i) and CP 3-2-III:4 (j) from family 3-2 with the S80fs mutation in MCTP2.

**Figure S7:**
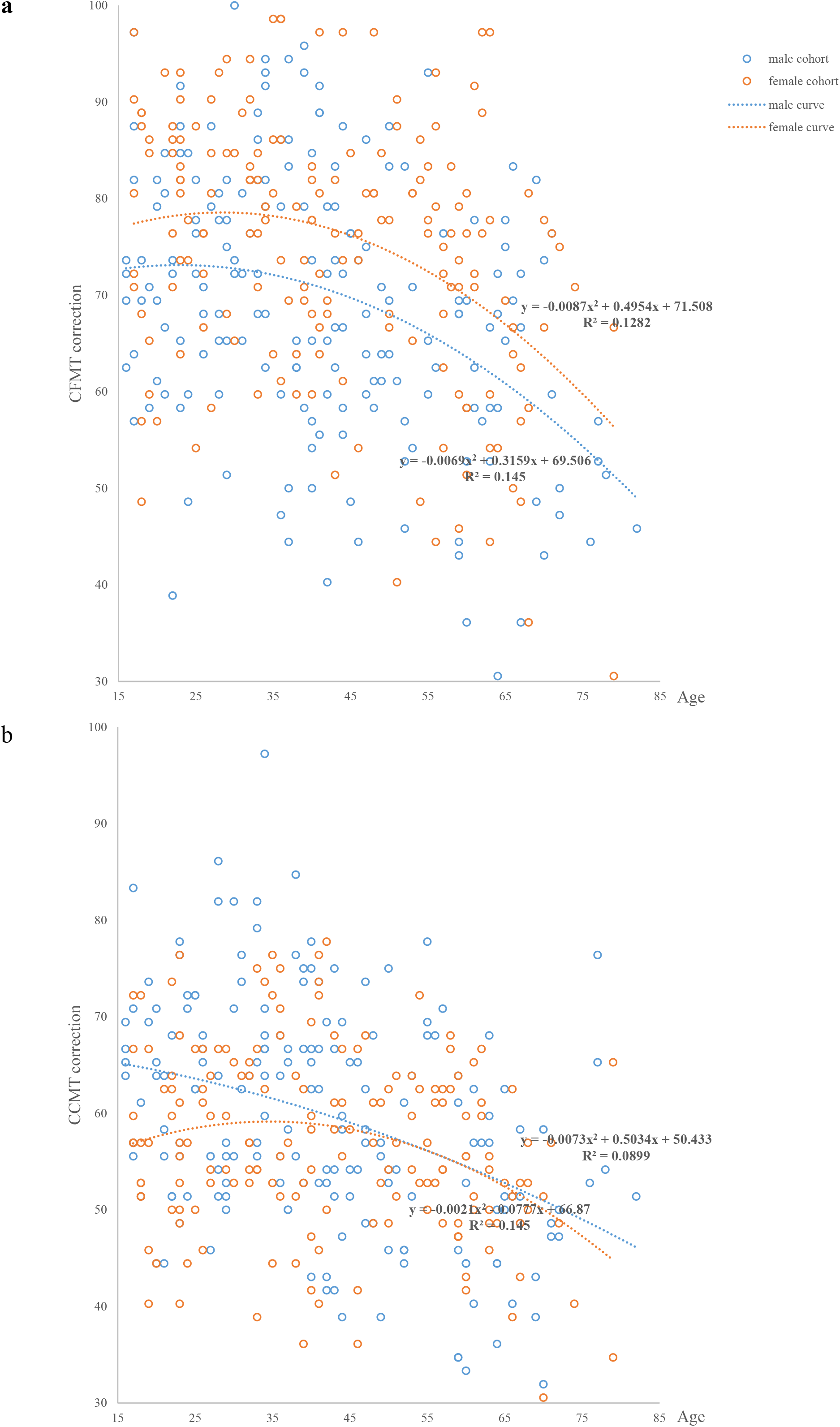
Scatterplots of Age against Total Correct for (a) the Cambridge Face Memory Test-Chinese (CFMT-C), and (b) the Cambridge Car Memory Test (CCMT) Male control individuals are shown as blue circles; female control individuals are shown as red circles. The curves show the best fitting second-order polynomial relating age to the relevant performance measure with different genders, blue for male, red for female.

**Figure S8:**
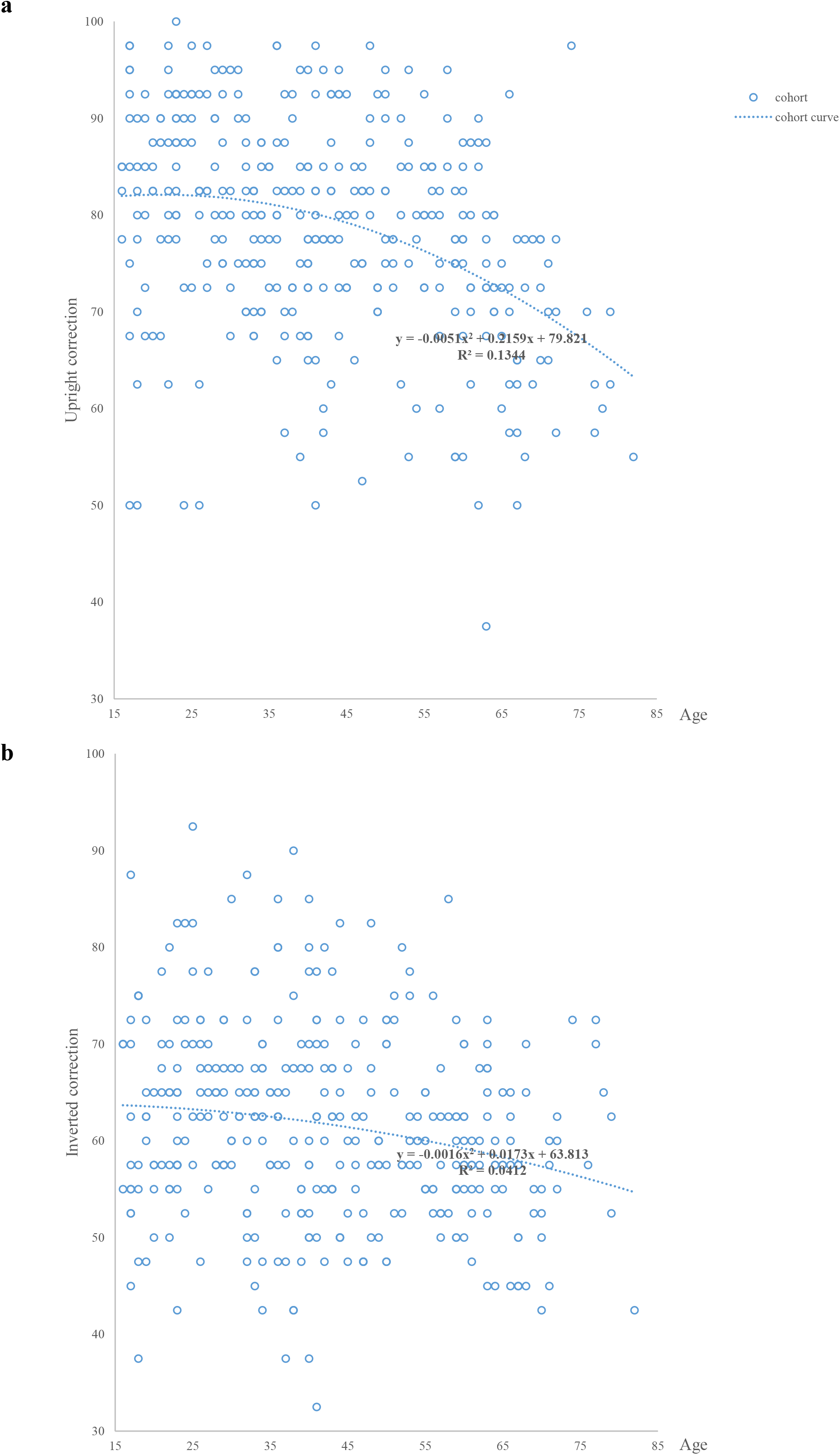
Scatterplots of Age against Total Correct for (a) the Upright Face, and (b) the Inverted Face. Control individuals are shown as blue circles. The curves show the best fitting second-order polynomial relating age to the relevant performance measure.

**Table S1.** Summary of Phenotypic Analysis and Genetic Analysis of Each Member of Family A.

| Family Member | obvious difficulty in face recognition in daily life by interview | impaired on at least one of the face behavioral tests | Linkage 1 | Linkage 2 | Array | WES | WGS | fMRI |
| --- | --- | --- | --- | --- | --- | --- | --- | --- |
|  |  |  | cases defined by interview and tests | cases defined by interview |  |  |  |  |
| IV:2 | V | - | CP | CP | V | - | - | - |
| IV:3 |  | - | normal | normal | V | - | - | - |
| IV:4 | V | - | CP | CP | V | - | - | - |
| IV:5 |  | - | normal | normal | V | - | - | - |
| IV:6 | V | - | CP | CP | V | - | - | - |
| IV:7 |  | - | normal | normal | V | - | - | - |
| IV:8 |  | - | normal | normal | V | - | - | - |
| IV:9 |  | - | normal | normal | V | - | - | - |
| M61 | V | - | CP | CP | V | - | - | - |
| IV:11 |  | - | normal | normal | V | - | - | - |
| V:1 |  | V | CP | CP | V | - | - | - |
| V:3 |  |  | normal | normal | V | - | - | - |
| V:4 | V | V | CP | CP | V | - | - | - |
| V:6 | V | V | CP | CP | V | V | - | - |
| V:7 |  |  | normal | normal | V | - | - | - |
| V:8 |  |  | normal | normal | V | - | - | - |
| V:9 | V | V | CP | CP | V | V | - | - |
| V:10 | V |  | unclear | CP | V | - | - | - |
| V:11 | V | V | CP | CP | V | V | - | - |
| V:12 |  |  | normal | normal | V | - | - | - |
| V:13 | V | V | CP | CP | V | - | - | - |
| V:14 |  |  | normal | normal | V | - | - | - |
| V:15 | V | V | CP | CP | V | - | - | - |
| V:16 |  |  | normal | normal | V | - | - | - |
| V:19 | V | V | CP | CP | V | V | V | - |
| VI:1 |  |  | normal | normal | V | - | - | V |
| VI:2 |  |  | normal | normal | V | - | - | V |
| VI:3 |  |  | normal | normal | V | - | - | - |
| VI:4 |  |  | normal | normal | V | - | - | - |
| VI:5 | V | V | CP | CP | V | - | V | V |
| VI:6 | V |  | unclear | CP | V | - | - | - |
| VI:7 | V |  | unclear | CP | V | - | - | V |
| VI:8 | V | V | CP | CP | V | - | - | - |
| VI:9 | V |  | - | - | - | - | - | V |
| VI:10 |  |  | normal | normal | V | - | - | V |

**Table S2.** Summary of Computer Simulations of Linkage Analysis.

|  | independent linked regions |  | observed maximum LOD score | empirical p value |
| --- | --- | --- | --- | --- |
| Linkage 1 | chr2 | 18150570-19775673 | 1.325 | 0.932 |
|  | chr6 | 102275330-103291012 | 1.192 | 0.942 |
|  | chr12 | 21989321-22891967 | 1.867 | 0.591 |
|  | chr14 | 26341293-29161732 | 1.657 | 0.659 |
|  | chr15 | 94266567-96809782 | 3.196 | **0.017** |
| Linkage 2 | chr10 | 110316481-112555472 | 1.826 | 0.528 |
|  | chr12 | 22015114-22325528 | 1.095 | 0.854 |
|  | chr15 | 94245722-97520230 | 4.270 | **0.009** |

**Table S3.**
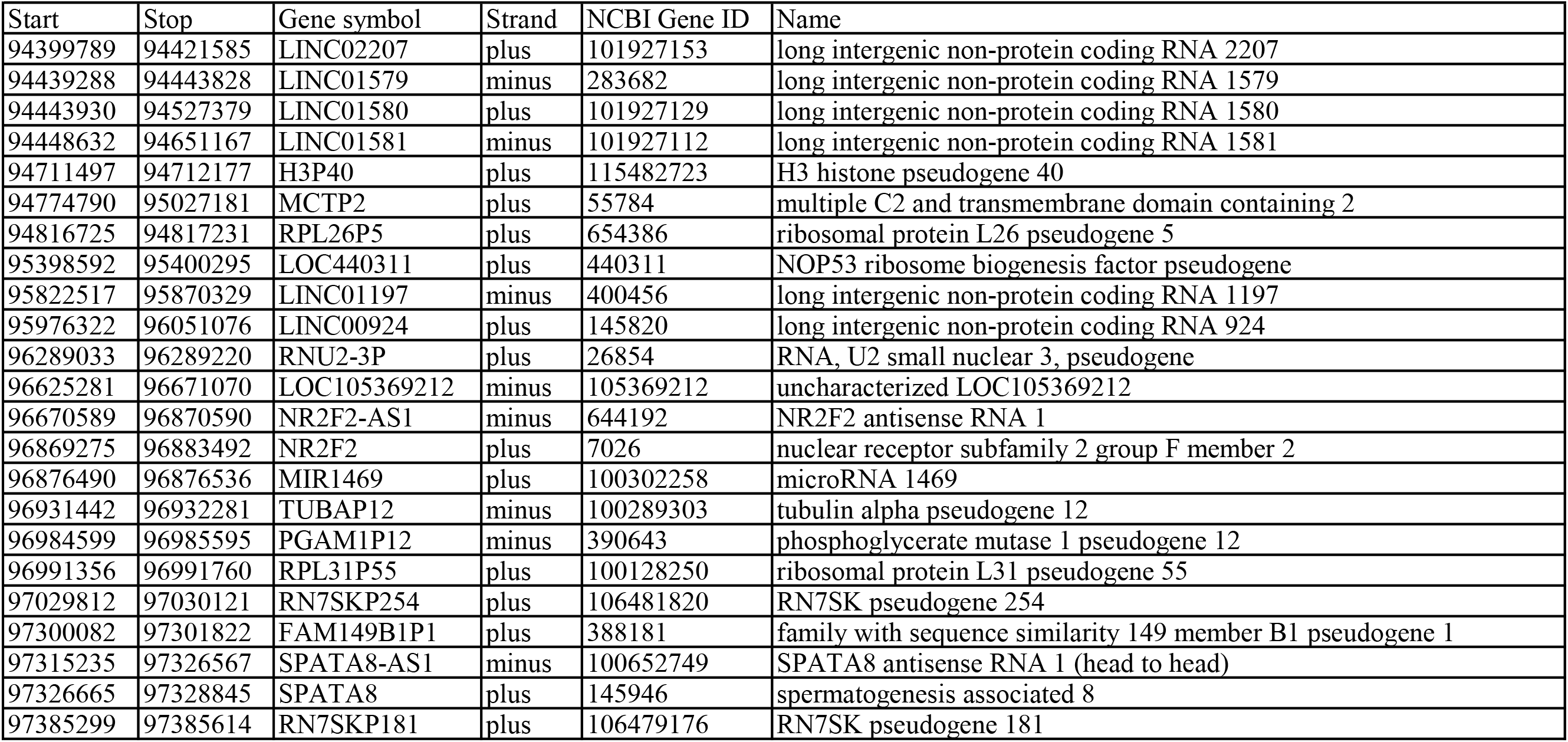
Candidate Genes Annotated by the NCBI Database (NC_000015.9) in Regions of Linkage Analyses with LOD Scores over 3.

| Start | Stop | Gene symbol | Strand | NCBI Gene ID | Name |
| --- | --- | --- | --- | --- | --- |
| 94399789 | 94421585 | LINC02207 | plus | 101927153 | long intergenic non-protein coding RNA 2207 |
| 94439288 | 94443828 | LINC01579 | minus | 283682 | long intergenic non-protein coding RNA 1579 |
| 94443930 | 94527379 | LINC01580 | plus | 101927129 | long intergenic non-protein coding RNA 1580 |
| 94448632 | 94651167 | LINC01581 | minus | 101927112 | long intergenic non-protein coding RNA 1581 |
| 94711497 | 94712177 | H3P40 | plus | 115482723 | H3 histone pseudogene 40 |
| 94774790 | 95027181 | MCTP2 | plus | 55784 | multiple C2 and transmembrane domain containing 2 |
| 94816725 | 94817231 | RPL26P5 | plus | 654386 | ribosomal protein L26 pseudogene 5 |
| 95398592 | 95400295 | LOC440311 | plus | 440311 | NOP53 ribosome biogenesis factor pseudogene |
| 95822517 | 95870329 | LINC01197 | minus | 400456 | long intergenic non-protein coding RNA 1197 |
| 95976322 | 96051076 | LINC00924 | plus | 145820 | long intergenic non-protein coding RNA 924 |
| 96289033 | 96289220 | RNU2-3P | plus | 26854 | RNA, U2 small nuclear 3, pseudogene |
| 96625281 | 96671070 | LOC105369212 | minus | 105369212 | uncharacterized LOC105369212 |
| 96670589 | 96870590 | NR2F2-AS1 | minus | 644192 | NR2F2 antisense RNA 1 |
| 96869275 | 96883492 | NR2F2 | plus | 7026 | nuclear receptor subfamily 2 group F member 2 |
| 96876490 | 96876536 | MIR1469 | plus | 100302258 | microRNA 1469 |
| 96931442 | 96932281 | TUBAP12 | minus | 100289303 | tubulin alpha pseudogene 12 |
| 96984599 | 96985595 | PGAM1P12 | minus | 390643 | phosphoglycerate mutase 1 pseudogene 12 |
| 96991356 | 96991760 | RPL31P55 | plus | 100128250 | ribosomal protein L31 pseudogene 55 |
| 97029812 | 97030121 | RN7SKP254 | plus | 106481820 | RN7SK pseudogene 254 |
| 97300082 | 97301822 | FAM149B1P1 | plus | 388181 | family with sequence similarity 149 member B1 pseudogene 1 |
| 97315235 | 97326567 | SPATA8-AS1 | minus | 100652749 | SPATA8 antisense RNA 1 (head to head) |
| 97326665 | 97328845 | SPATA8 | plus | 145946 | spermatogenesis associated 8 |
| 97385299 | 97385614 | RN7SKP181 | plus | 106479176 | RN7SK pseudogene 181 |

**Table S4.** the Summary Analysis of the Sequencing Data.

| Terms | Exome<br>Sequence of<br>Family A | Whole Genome<br>Sequence of<br>Family A | Candidate Region Sequence<br>of the second cohort of 1928<br>medical students | Candidate Region Sequence<br>of the third cohort of 1757<br>medical students |
| --- | --- | --- | --- | --- |
| Sample number | 4 | 2 | 1928 | 1757 |
| Reads_mapped (%) | 99.86 | 99.86 | 99.91 | 98.99 |
| Target_Efficiency (%) | 83.48 | - | 72.48 | 82.57 |
| Mean Depth | 83.55 | 42.66 | 10255 | 4501.25 |
| >=10Coverage (%) | 94.6 | 95.55 | 95.44 | 92.24 |
| >=20Coverage (%) | 89.2 | 90.05 | 94.40 | 88.58 |
| >=40Coverage (%) | 77.1 | 56.7 | 93.13 | 83.84 |
| >=100Coverage (%) | 33.3 | - | 90.57 | 76.29 |

**Table S5.**
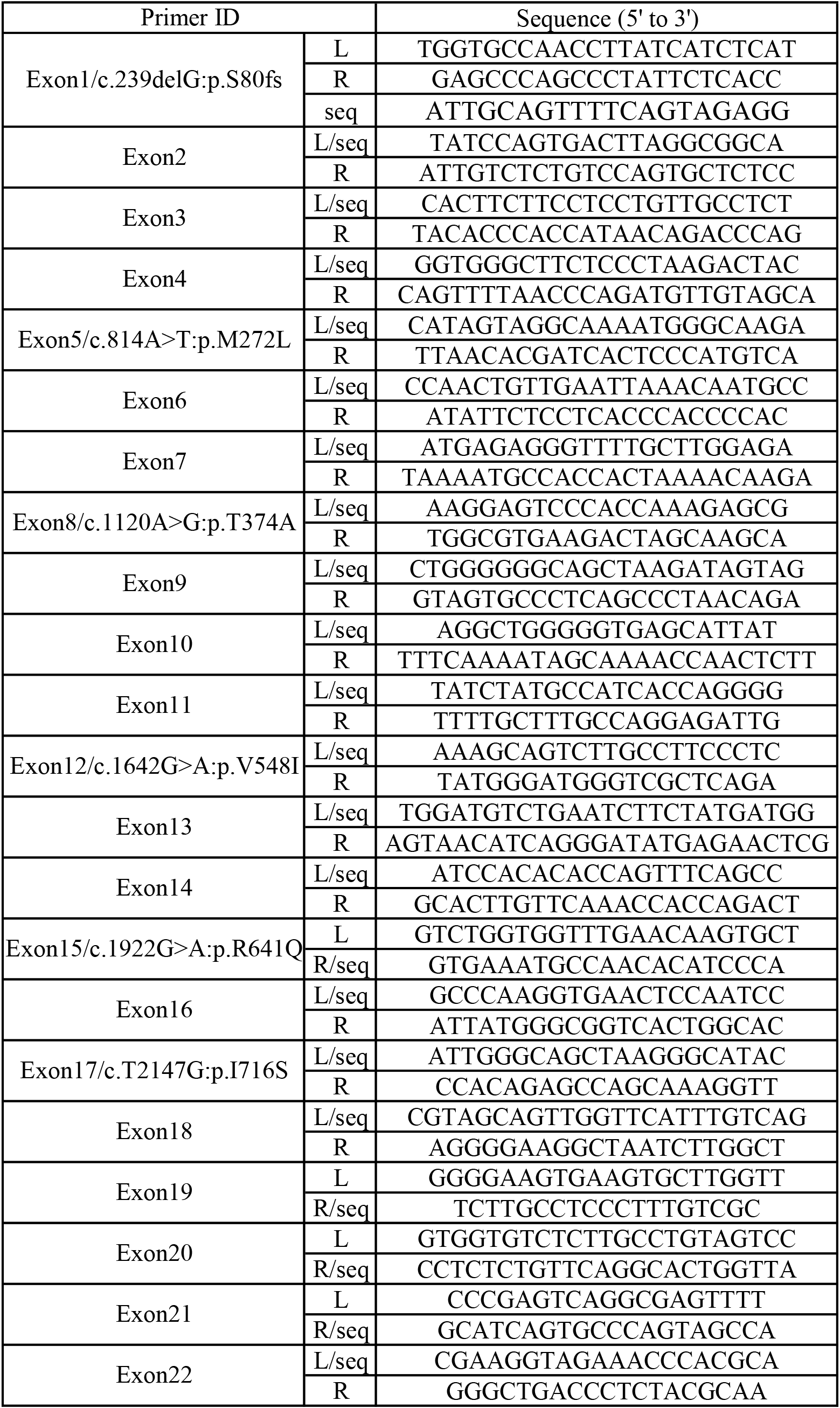
PCR Primer Sequences for Each Exon and Mutation Site of MCTP2. L, the left primer; R, the right primer; seq, the primer for sequencing

**Table S6.**
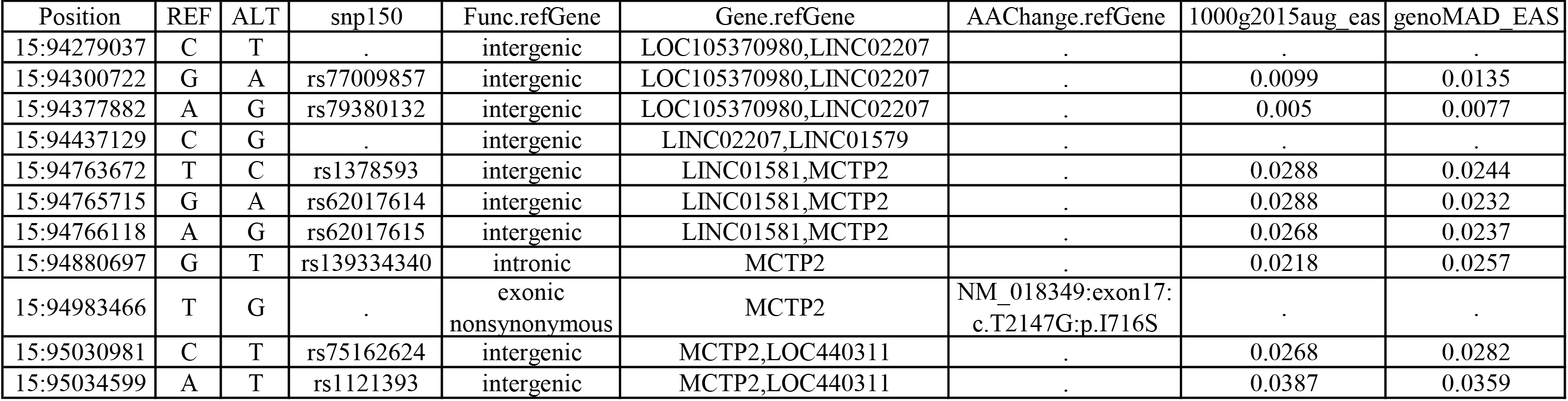
11 Heterozygous Variants with MAF under 0.05 Shared with the Affected Individuals of Family A by WGS.

| Position | REF | ALT | snp150 | Func.refGene | Gene.refGene | AACChange.refGene | 1000g2015aug_eas | genoMAD_EAS |
| --- | --- | --- | --- | --- | --- | --- | --- | --- |
| 15:94279037 | C | T | . | intergenic | LOC105370980,LINC02207 | . | . | . |
| 15:94300722 | G | A | rs77009857 | intergenic | LOC105370980,LINC02207 | . | 0.0099 | 0.0135 |
| 15:94377882 | A | G | rs79380132 | intergenic | LOC105370980,LINC02207 | . | 0.005 | 0.0077 |
| 15:94437129 | C | G | . | intergenic | LINC02207,LINC01579 | . | . | . |
| 15:94763672 | T | C | rs1378593 | intergenic | LINC01581,MCTP2 | . | 0.0288 | 0.0244 |
| 15:94765715 | G | A | rs62017614 | intergenic | LINC01581,MCTP2 | . | 0.0288 | 0.0232 |
| 15:94766118 | A | G | rs62017615 | intergenic | LINC01581,MCTP2 | . | 0.0268 | 0.0237 |
| 15:94880697 | G | T | rs139334340 | intronic | MCTP2 | . | 0.0218 | 0.0257 |
| 15:94983466 | T | G | . | exonic<br>nonsynonymous | MCTP2 | NM_018349:exon17:<br>c.T2147G:p.I716S | . | . |
| 15:95030981 | C | T | rs75162624 | intergenic | MCTP2,LOC440311 | . | 0.0268 | 0.0282 |
| 15:95034599 | A | T | rs1121393 | intergenic | MCTP2,LOC440311 | . | 0.0387 | 0.0359 |

**Table S7.**
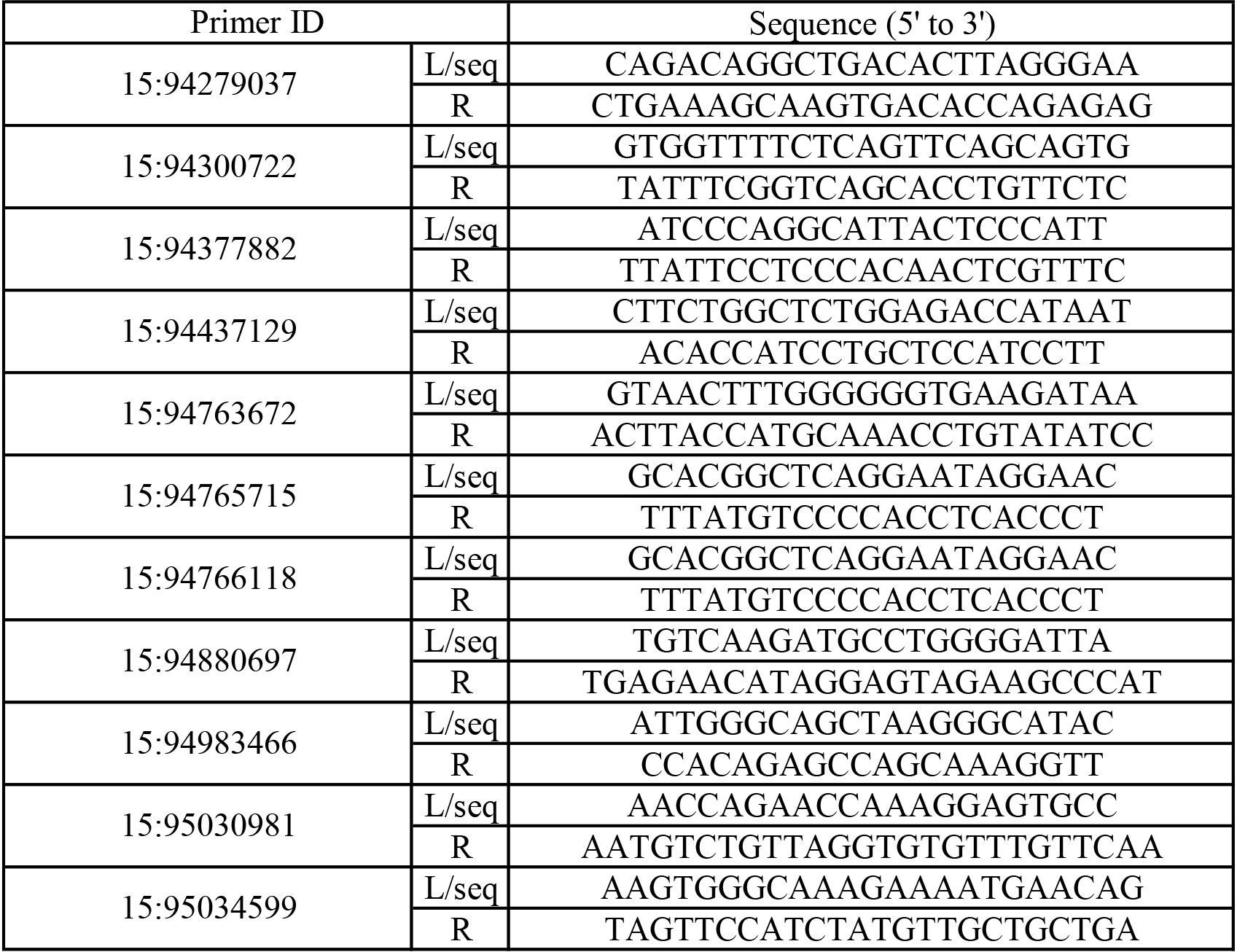
PCR Primer Sequences for 11 Heterozygous Variants (MAF under 0.05) Shared by the Affected Individuals of Family A by WGS.

**Table S8.** MCTP2 Mutations Found in Sporadic CP Candidates Screened by a 20-Item Questionnaire. NS: nonsynonymous single nucleotide variant; FD: frameshift_deletion; AAF_EAS: alternative allele frequency in EAS (East Asian) population; SIFT: predicts whether an amino acid substitution affects protein function as D (Deleterious) or T (tolerated) by SIFT algorithm; Polyphen2_HDIV, Polyphen2_HVAR: predicts possible impact of an amino acid substitution on the structure and function of a human protein using straightforward physical and comparative considerations as D (Probably damaging), P (possibly damaging) or B (benign) by PolyPhen 2 (Polymorphism Phenotyping v2); LRT: uses comparative genomics to predict whether variants disrupt highly conserved amino acids as D (deleterious), N (neutral) or U (unknown) by LRT (Likelihood Ratio Test); Mutation Taster: predicts an alteration as one of four possible types: D (disease causing - i.e. probably deleterious), A (disease causing automatic - i.e. known to be deleterious), N (polymorphism - i.e. probably harmless), P (polymorphism automatic - i.e. known to be harmless). These abbreviations also apply to the following tables. * Start Position Based on the MCTP2 RefSeq (hg19, NM_018349); # dbSNP build 150, & RefGene: NM_018349.

| Patients | Chr15 Position* | RefSNP ID# | Type of Variants | Amino Acid Change & | AAF_EAS |  |  |  | Non-Synonymous Variants Annotation |  |  |  |  |
| --- | --- | --- | --- | --- | --- | --- | --- | --- | --- | --- | --- | --- | --- |
|  |  |  |  |  | 1000G | ExAC | gnomAD |  | SIFT | Polyphen2 HDIV | Polyphen2 HVAR | LRT | Mutation Taster |
|  |  |  |  |  |  |  | exome | exome |  |  |  |  |  |
| 1-1<br>1-2<br>1-3 | 94841733 | rs574533536 | FD | exon1:c.239delG;p.S80fs | 0.005 | 0.0031 | 0.0036 | 0.0045 | . | . | . | . | . |
| 1-4 | 94883461 | rs773393905 | NS | exon5:c.A814T;p.M272L | . | 0.0001 | . | . | T | P | P | D | D |
| 1-5 | 94899480 | rs764700909 | NS | exon8:c.A1120G;p.T374A | . | 0.0002 | 0.0002 | . | T | B | B | N | D |
| 1-6 | 94927310 | rs182595113 | NS | exon12:c.G1642A;p.V548I | 0.003 | 0.0008 | 0.0013 | 0.0006 | T | B | B | N | N |
| 1-7 | 94943181 | rs749217960 | NS | exon15:c.G1922A;p.R641Q | . | . | 0.0000544 | . | D | P | B | D | D |

**Table S9.**
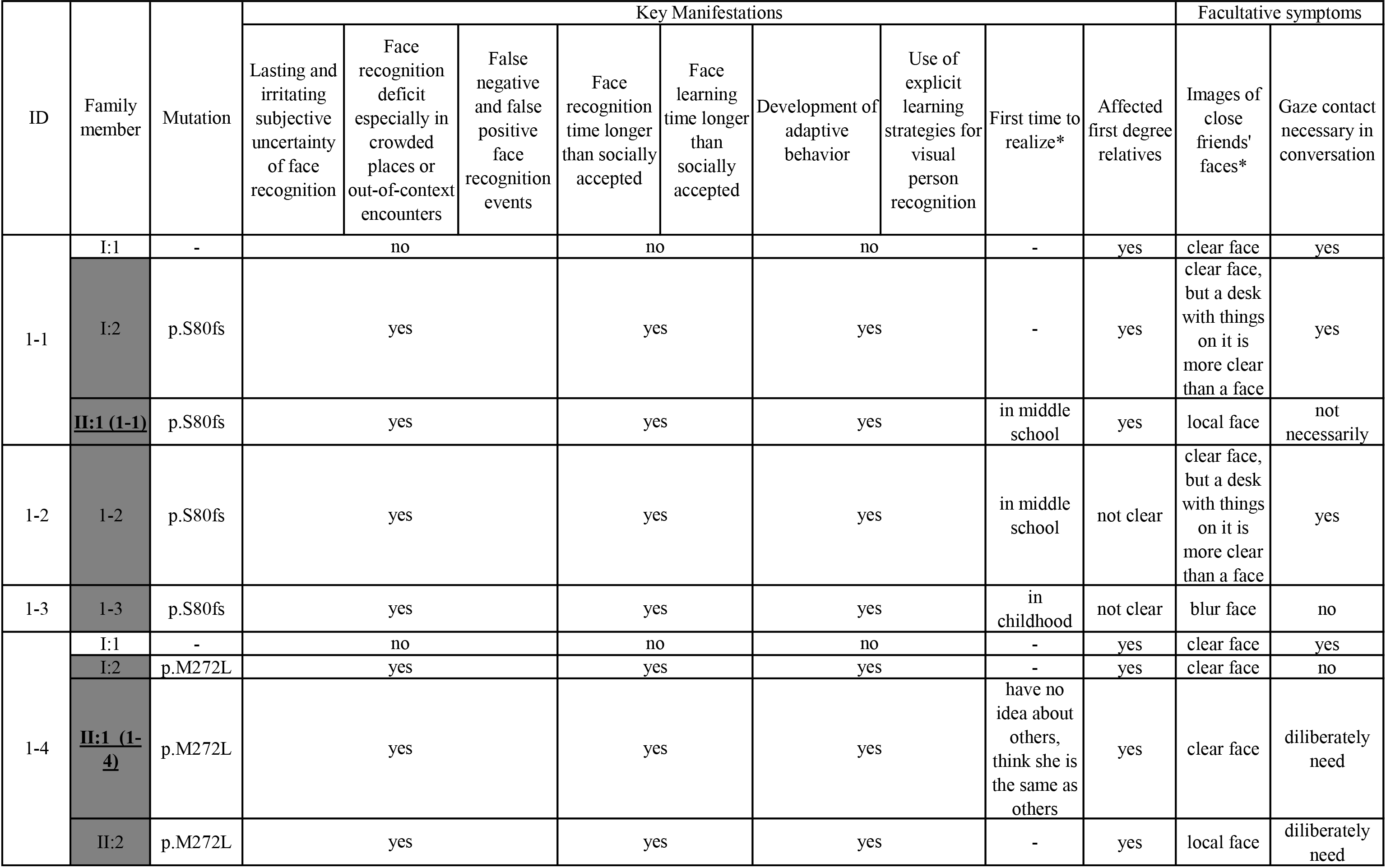
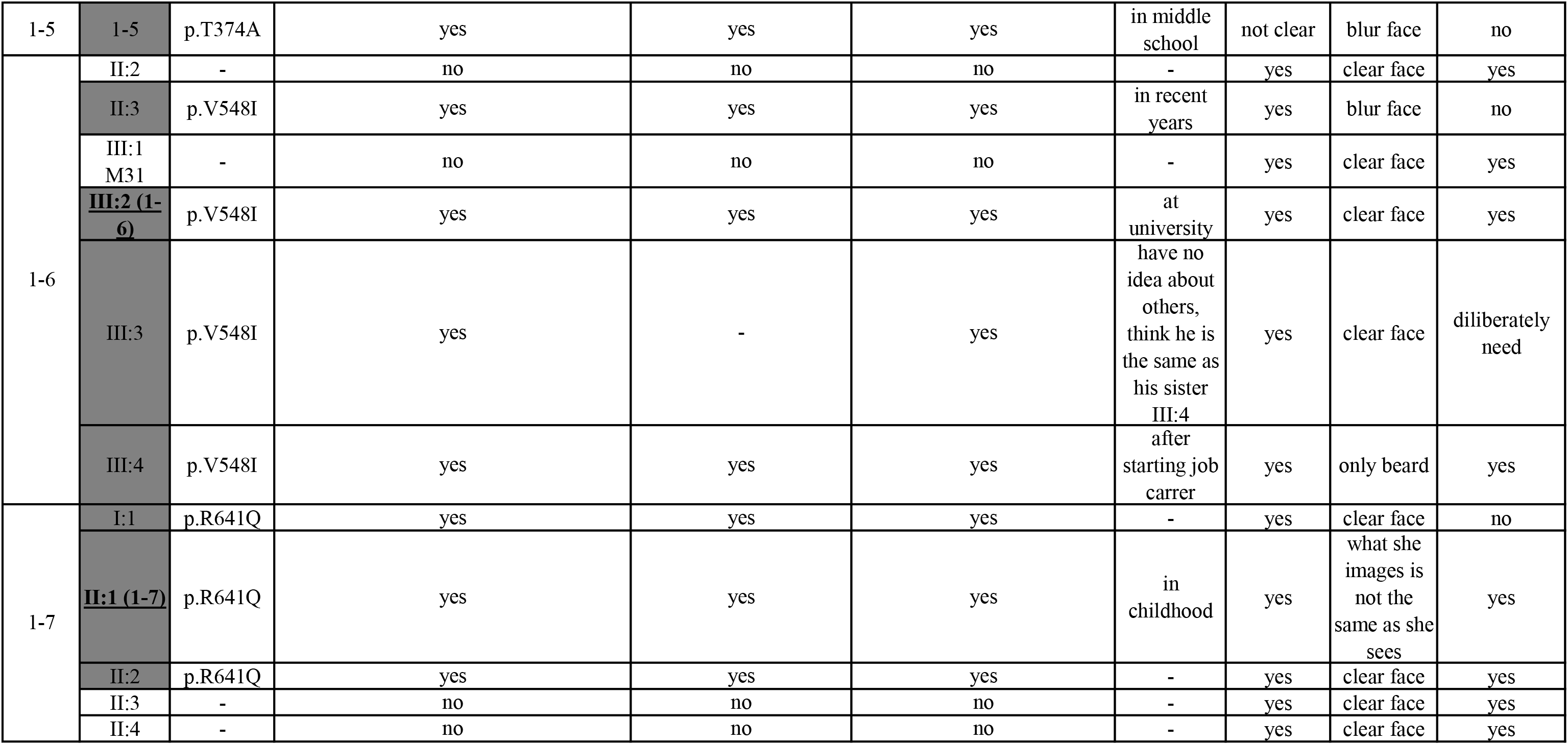
Most Discriminative Symptoms for the Diagnosis of Additional CP Individuals and Families. Member codes with dark grey background indicate CP candidates who showed daily hallmarks of CP. The index for each family is shown in bold and underlined. Whether there is an obvious key manifestation for CP is expressed by yes or no. More detailed diagnostic information can be obtained by contacting the corresponding author.

| ID | Family member | Mutation | Key Manifestations |  |  |  |  |  |  |  |  | Facultative symptoms |  |
| --- | --- | --- | --- | --- | --- | --- | --- | --- | --- | --- | --- | --- | --- |
|  |  |  | Lasting and irritating subjective uncertainty of face recognition | Face recognition deficit especially in crowded places or out-of-context encounters | False negative and false positive face recognition events | Face recognition time longer than socially accepted | Face learning time longer than socially accepted | Development of adaptive behavior | Use of explicit learning strategies for visual person recognition | First time to realize* | Affected first degree relatives | Images of close friends' faces* | Gaze contact necessary in conversation |
| 1-1 | I:1 | - | no |  |  | no |  | no |  | - | yes | clear face | yes |
|  | I:2 | p.S80fs | yes |  |  | yes |  | yes |  | - | yes | clear face, but a desk with things on it is more clear than a face | yes |
|  | <u>II:1 (1-1)</u> | p.S80fs | yes |  |  | yes |  | yes |  | in middle school | yes | local face | not necessarily |
| 1-2 | 1-2 | p.S80fs | yes |  |  | yes |  | yes |  | in middle school | not clear | clear face, but a desk with things on it is more clear than a face | yes |
| 1-3 | 1-3 | p.S80fs | yes |  |  | yes |  | yes |  | in childhood | not clear | blur face | no |
| 1-4 | I:1 | - | no |  |  | no |  | no |  | - | yes | clear face | yes |
|  | I:2 | p.M272L | yes |  |  | yes |  | yes |  | - | yes | clear face | no |
|  | <u>II:1 (1-4)</u> | p.M272L | yes |  |  | yes |  | yes |  | have no idea about others, think she is the same as others | yes | clear face | diliberately need |
|  | II:2 | p.M272L | yes |  |  | yes |  | yes |  | - | yes | local face | diliberately need |

|  |  |  |  |  |  |  |  |  |  |
| --- | --- | --- | --- | --- | --- | --- | --- | --- | --- |
| 1-5 | 1-5 | p.T374A | yes | yes | yes | in middle school | not clear | blur face | no |
| 1-6 | II:2 | - | no | no | no | - | yes | clear face | yes |
|  | II:3 | p.V548I | yes | yes | yes | in recent years | yes | blur face | no |
|  | III:1 M31 | - | no | no | no | - | yes | clear face | yes |
|  | **<u>III:2 (1-6)</u>** | p.V548I | yes | yes | yes | at university | yes | clear face | yes |
|  | III:3 | p.V548I | yes | - | yes | have no idea about others, think he is the same as his sister III:4 | yes | clear face | diliberately need |
|  | III:4 | p.V548I | yes | yes | yes | after starting job carrer | yes | only beard | yes |
| 1-7 | I:1 | p.R641Q | yes | yes | yes | - | yes | clear face | no |
|  | **<u>II:1 (1-7)</u>** | p.R641Q | yes | yes | yes | in childhood | yes | what she images is not the same as she sees | yes |
|  | II:2 | p.R641Q | yes | yes | yes | - | yes | clear face | yes |
|  | II:3 | - | no | no | no | - | yes | clear face | yes |
|  | II:4 | - | no | no | no | - | yes | clear face | yes |

**Table S10.** Rare Variants (MAF<0.005) in MCTP2 from a Second Cohort of 1928 Students.

| Chr15<br>Position* | RefSNP ID# | AA Change.refGene | AAF |  |  |  | Non-Synonymous Variants Annotation |  |  |  |  |
| --- | --- | --- | --- | --- | --- | --- | --- | --- | --- | --- | --- |
|  |  | NM_018349 | 1000G2015A<br>ug_EAS | ExAC_EAS | gnomAD_<br>EAS | our cohort of<br>1928<br>individuals | SIFT | Polyphen2<br>HDIV | Polyphen2<br>HVAR | LRT | Mutation<br>Taster |
| 94841499 | rs776199245 | exon1:c.A5T:p.D2V | . | 0.0004 | 0.0006 | 0.0003 | D | D | D | D | D |
| 94841634 | rs149133063 | exon1:c.G140A:p.R47H | . | 0.0001 | 0 | 0.0003 | D | D | P | D | D |
| 94841676 | . | exon1:c.T182A:p.L61X | . | . | . | 0.0003 | . | . | . | D | A |
| 94841690 | rs141454907 | exon1:c.C196T:p.R66W | . | 0.0002 | 0.0006 | 0.0003 | D | D | B | N | N |
| 94841732 | rs574533536 | exon1:c.239delG:p.S80Tfs*17 | 0.005 | 0.0031 | 0.0045 | 0.0026 | . | . | . | . | . |
| 94841776 | rs376018109 | exon1:c.C282G:p.S94R | . | 0.0009 | 0.0039 | 0.0010 | D | P | B | N | D |
| 94841843 | rs748435445 | exon1:c.349_350insAAACAGA:p.<br>D119Efs*13 | . | . | . | 0.0003 | . | . | . | . | . |
| 94841861 | rs200962853 | exon1:c.G367A:p.A123T | 0.004 | 0.0045 | 0.0045 | 0.0075 | T | B | B | N | N |
| 94841867 | rs377631883 | exon1:c.G373A:p.A125T | . | 0.0001 | 0 | 0.0005 | T | B | B | N | N |
| 94841889 | rs199860245 | exon1:c.G395A:p.R132Q | 0.002 | 0.0011 | 0.0019 | 0.0023 | T | B | B | N | N |
| 94841934 | rs764405023 | exon1:c.G440T:p.G147V | . | 0.0005 | 0.0013 | 0.0016 | D | P | B | N | N |
| 94857525 | rs749409827 | exon2:c.C480A:p.S160R | . | 0.0001 | . | 0.0005 | D | P | B | N | N |
| 94857550 | rs768863067 | exon2:c.T505C:p.S169P | . | 0.0005 | . | 0.0003 | D | D | D | D | D |
| 94857572 | rs772412690 | exon2:c.C527G:p.S176C | . | 0.0004 | . | 0.0013 | T | B | B | N | D |
| 94858867 | rs750195259 | exon3:c.637+1->T | . | 0.0016 | 0.0006 | 0.0013 | . | . | . | . | . |
| 94882518 | rs756489641 | exon4:c.638-1G>C | . | 0.0001 | . | 0.0005 | . | . | . | . | D |
| 94882549 | . | exon4:c.T668C:p.L223P | . | . | . | 0.0003 | D | D | D | D | D |
| 94883462 | . | exon5:c.T815C:p.M272T | . | . | . | 0.0003 | D | D | D | D | D |
| 94884094 | rs199933047 | exon6:c.A910C:p.M304L | 0.001 | . | 0.0006 | 0.0003 | T | P | B | D | D |
| 94884106 | rs145490381 | exon6:c.G922C:p.V308L | 0.002 | . | 0.0006 | 0.0003 | T | B | B | D | D |
| 94899388 | rs77311454 | exon8:c.G1028A:p.R343Q | . | 0.0001 | 0 | 0.0005 | D | D | P | D | D |
| 94899411 | rs762149227 | exon8:c.A1051G:p.N351D | . | 0.0001 | . | 0.0005 | D | D | P | D | D |
| 94899432 | rs199572394 | exon8:c.A1072G:p.I358V | . | . | . | 0.0003 | T | B | B | D | D |
| 94899475 | . | exon8:c.G1115A:p.S372N | . | . | . | 0.0003 | T | B | B | N | D |
| 94899480 | rs764700909 | exon8:c.A1120G:p.T374A | . | 0.0002 | . | 0.0005 | T | B | B | N | D |
| 94899488 | . | exon8:c.G1128A:p.M376I | . | . | . | 0.0003 | T | B | B | N | D |
| 94899522 | . | exon8:c.A1162G:p.K388E | . | . | . | 0.0003 | D | D | D | D | D |
| 94901803 | rs199614826 | exon9:c.G1263A:p.W421X | 0.001 | 0.0006 | . | 0.0005 | . | . | . | D | A |
| 94901805 | rs761523056 | exon9:c.G1265A:p.G422E | . | 0.0002 | . | 0.0003 | T | B | B | N | D |
| 94901834 | rs747502432 | exon9:c.1295delT:p.L432Rfs*13 | . | 0.0006 | . | 0.0026 | . | . | . | . | . |
| 94901842 | rs746093287 | exon9:c.1301+1G>A | . | 0 | . | 0.0005 | . | . | . | . | D |
| 94910856 | . | exon10:c.C1324T:p.L442F | . | . | . | 0.0003 | T | D | P | D | D |
| 94910913 | . | exon10:c.C1381T:p.L461F | . | . | . | 0.0005 | D | D | D | D | N |
| 94910933 | . | exon10:c.1402dupA:p.T468Nfs*10 | . | . | . | 0.0003 | . | . | . | . | . |
| 94911020 | rs2289010 | exon10:c.T1488A:p.Y496X | 0.001 | 0.0009 | 0.0006 | 0.0013 | . | . | . | D | A |
| 94913325 | . | exon11:c.A1498T:p.N500Y | . | . | . | 0.0003 | D | D | P | D | D |
| 94913349 | rs772110471 | exon11:c.G1522A:p.V508I | . | 0.0001 | . | 0.0005 | T | B | B | N | N |
| 94927287 | . | exon12:c.A1619G:p.N540S | . | . | . | 0.0003 | D | B | B | D | D |
| 94927292 | rs374061496 | exon12:c.C1624T:p.R542X | 0.004 | 0.0024 | 0.0032 | 0.0023 | . | . | . | D | A |
| 94927310 | rs182595113 | exon12:c.G1642A:p.V548I | 0.003 | 0.0008 | 0.0006 | 0.0003 | T | B | B | N | N |
| 94927336 | . | exon12:c.G1668A:p.W556X | . | . | . | 0.0003 | . | . | . | D | A |
| 94927338 | . | exon12:c.1671delC:p.N557Kfs*17 | . | . | . | 0.0003 | . | . | . | . | . |
| 94928755 | rs200626556 | exon13:c.1788+1G | 0.001 | 0.0005 | 0 | 0.0010 | . | . | . | . | D |
| 94942206 | rs138009671 | exon14:c.C1805T:p.P602L | . | 0.0001 | 0.0006 | 0.0008 | T | B | B | N | D |
| 94942260 | rs749725540 | exon14:c.T1859C:p.I620T | . | 0.0002 | . | 0.0003 | D | D | D | D | D |
| 94943181 | rs749217960 | exon15:c.G1922A:p.R641Q | . | 0 | . | 0.0003 | D | B | B | D | D |
| 94943208 | rs137927917 | exon15:c.G1949A:p.R650H | . | . | 0 | 0.0003 | T | D | P | D | D |
| 94945175 | . | exon16:c.T2012C:p.I671T | . | . | . | 0.0003 | D | B | B | D | D |
| 94945235 | rs576073379 | exon16:c.C2072T:p.T691I | 0.001 | 0.001 | 0.0013 | 0.0003 | T | B | B | N | N |
| 94945247 | rs370153540 | exon16:c.C2084T:p.A695V | . | 0.001 | 0.0006 | 0.0008 | T | B | B | N | N |
| 94983525 | rs751758425 | exon17:c.C2206T:p.Q736X | . | 0.0001 | . | 0.0005 | . | . | . | N | A |
| 94986160 | rs767710235 | exon18:c.A2221G:p.I741V | . | 0.0002 | 0.0006 | 0.0005 | T | B | B | N | N |
| 94986169 | . | exon18:c.G2230A:p.E744K | . | . | . | 0.0003 | D | B | B | D | D |
| 95019951 | rs889873034 | exon21:c.C2497T:p.R833X | . | . | 0.0006 | 0.0008 | . | . | . | D | A |
| 95019952 | rs187187495 | exon21:c.G2498A:p.R833Q | . | 0.0001 | 0 | 0.0003 | D | D | D | D | D |
| 95019969 | rs140817032 | exon21:c.G2515A:p.D839N | . | 0 | 0 | 0.0003 | T | D | D | D | D |
| 95022248 | rs746937797 | exon22:c.G2622T:p.K874N | . | 0.0002 | . | 0.0005 | D | D | D | N | D |

Note:\* Start Position Based on the MCTP2 RefSeq (hg19, NM\_018349);# dbSNP build 150

**Table S11.**
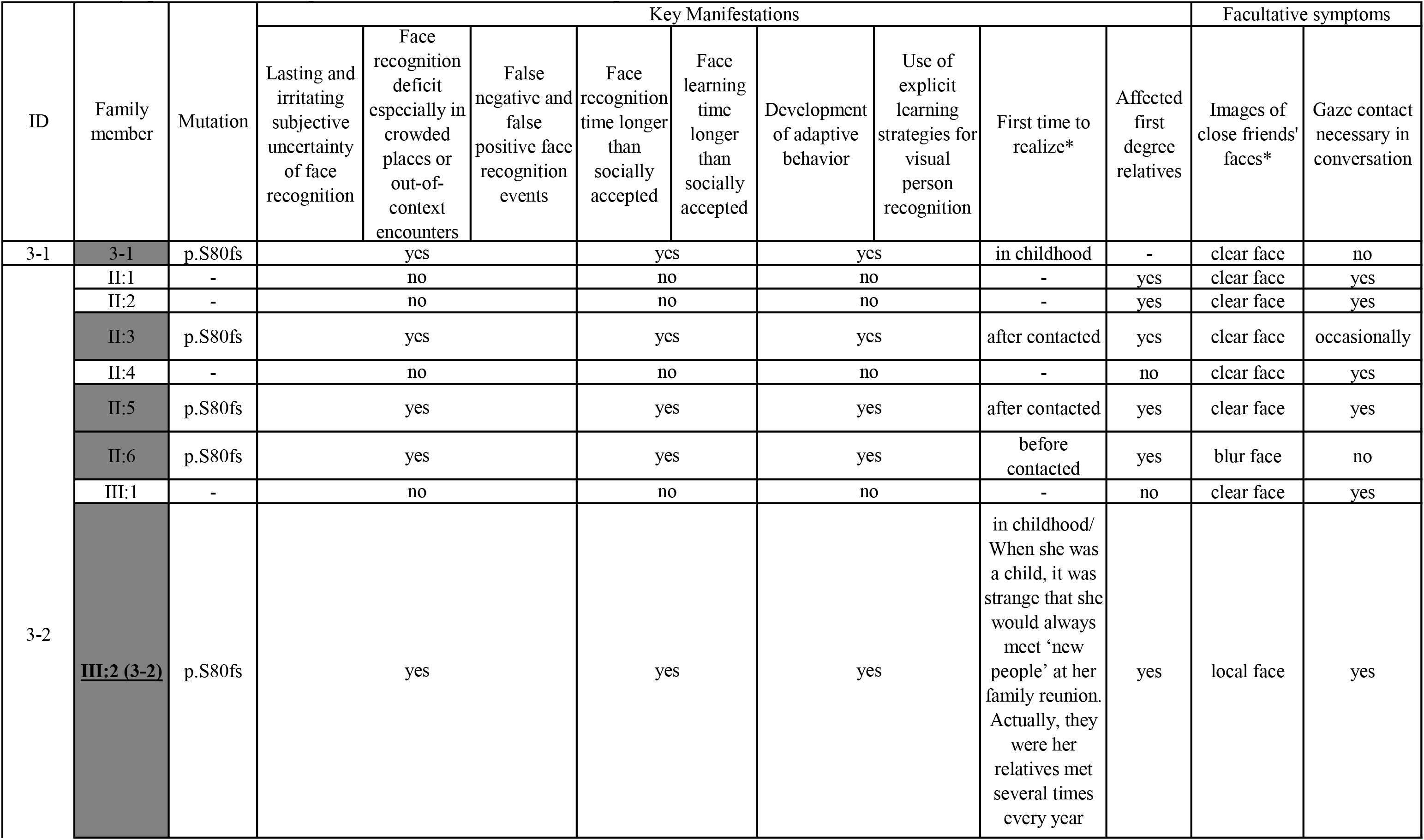
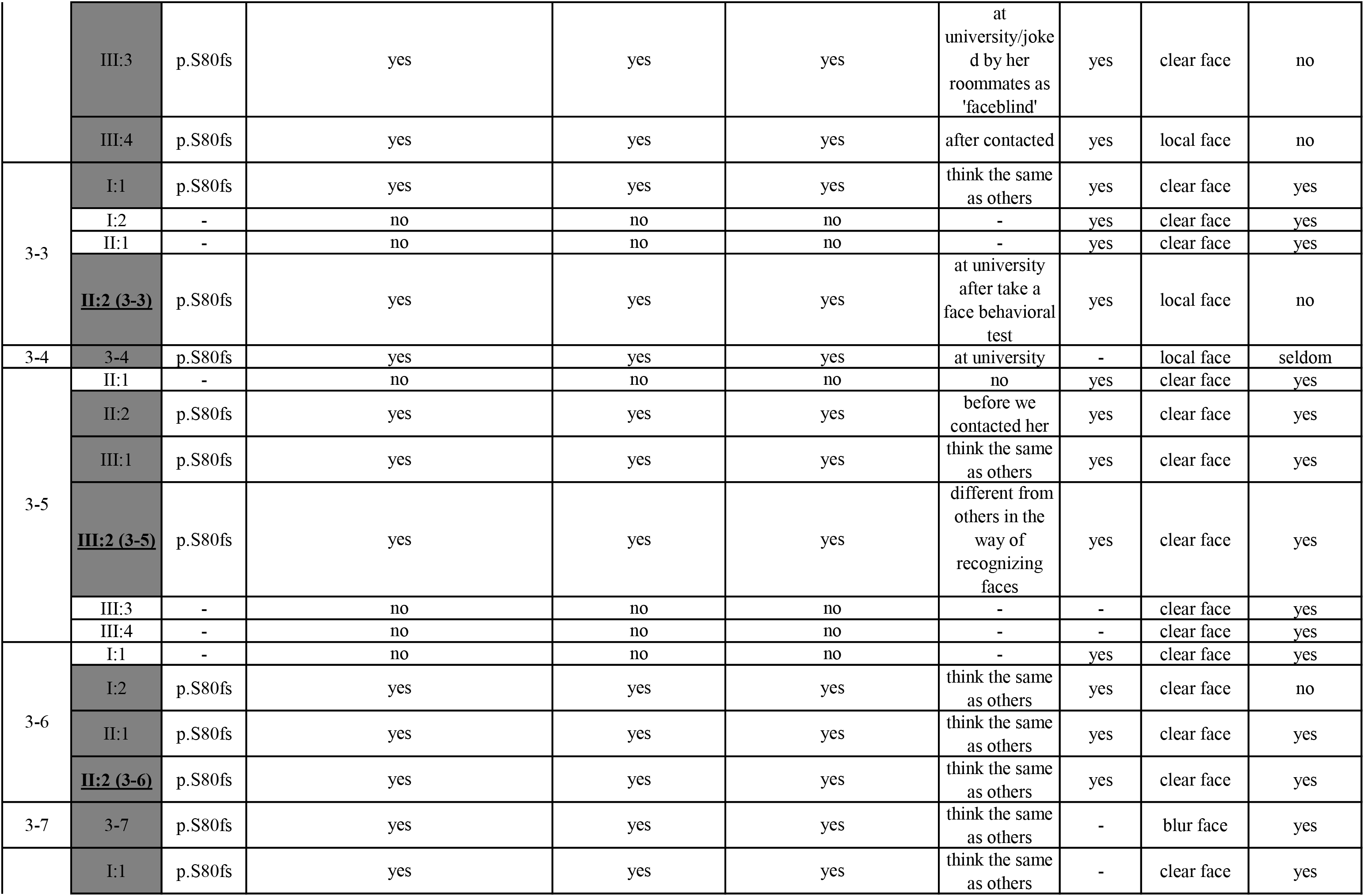
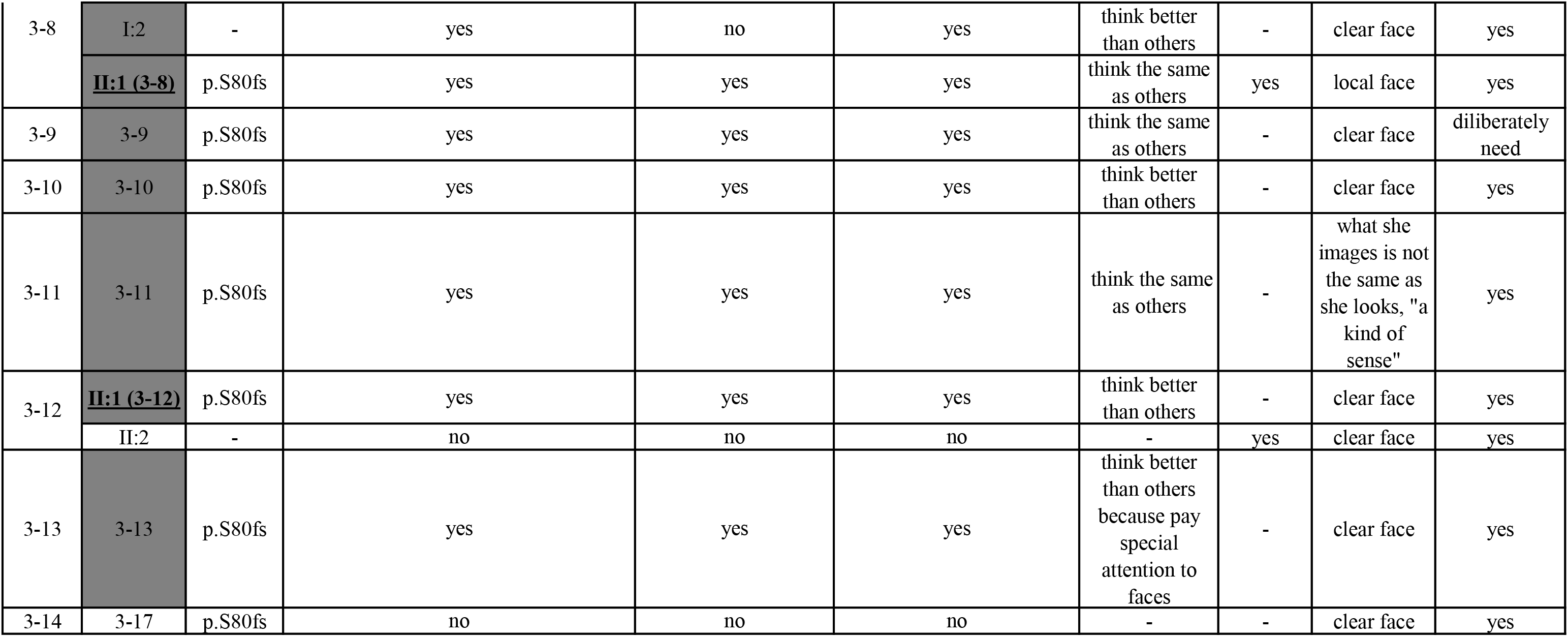
Symptoms for the Diagnosis in Persons with c.239delG, p.S80fs. Member codes with dark grey background indicate CP candidates who showed daily symptoms of CP. The index for each family is shown in bold and underlined. Whether there is an obvious key manifestation for CP is expressed by yes or no. More detailed diagnostic information can be obtained by contacting the corresponding author.

